# Large language model scoring of medical student reflection essays: Accuracy and reproducibility of prompt-model variations

**DOI:** 10.64898/2026.03.20.26348918

**Authors:** David A. Cook, Torrey A. Laack, V. Shane Pankratz

## Abstract

**Introduction:** Evaluate large language models (LLMs) for scoring medical student essays, and compare various prompting techniques and models.

**Methods:** OpenAI GPT scored 51 medical student reflection essays (15 real, 36 fabricated) using a previously-reported 6-point rubric (April-May 2025). We compared 29 prompt-model conditions by systematically varying the LLM prompts (including the persona, scoring rubric, few-shot learning [exemplars], chain-of-thought reasoning, and temperature), fine-tuning, and model (including GPT-4.1, GPT-4.1-mini, GPT-o4-mini, and GPT-4-Turbo). Outcomes were accuracy (compared with human raters, measured using single-score intraclass correlation coefficient [ICC] and mean absolute difference [MAD; zero indicates perfect agreement]), within-condition reproducibility, and cost.

**Results:** Across all conditions, it took mean (SD) 3.73 (3.12) seconds to score 1 essay. The cost to score 100 essays was USD $0.04 for GPT-4.1-mini, $0.21 for GPT-4.1, $0.57 for GPT-4.1 with 3 exemplars, and $2.00 for fine-tuned GPT-4.1. When the one-time cost of fine-tuning was amortized across 10,000 essays, the cost for fine-tuned GPT-4.1 was $0.20 per 100. Accuracy was “almost perfect” (ICC >0.80) for 28/29 conditions (97%). Fine-tuned models were more accurate than non-fine-tuned models (MAD difference –0.24 [95% CI, –0.34, –0.14]). Conditions with exemplars were more accurate than those without (MAD difference –0.44 [CI, –0.57, –0.31]). Accuracy progressively decreased as 6, 3, 1, and 0 rubric levels were explicitly defined in the prompt (P<.001). Contrary to hypotheses, accuracies for chain-of-thought prompts and variations in temperature and persona were not significantly different from the baseline prompt. Reproducibility ICC was >0.80 for 28/29 conditions (97%).

**Discussion:** Automated LLM essay scoring demonstrated near-perfect accuracy and reproducibility for most prompt-model conditions. Fine-tuned models and prompts with exemplars had higher accuracy but higher cost. Fine-tuned models had lower per-essay costs for larger essay volumes. For smaller volumes, non-fine-tuned GPT-4.1 provided excellent results at moderate cost. GPT-4.1-mini provided very good results at low cost.

## Introduction

Assessment drives learning, yet practical issues commonly constrain assessments to easily-quantifiable aspects of performance (e.g., multiple-choice assessments of knowledge).^1–4^ Qualitative (narrative) data, including “constructed response” assessments such as encounter notes and essays, offer a rich source of information appraising a broad range of learner attributes.^5–10^ However, scoring narrative assessments usually requires human raters. Implementing this at scale introduces practical challenges (e.g., rater availability, cost, and training) and scoring inconsistencies (rater fatigue, leniency/stringency, and random errors).^1, 11^ Computer automated scoring offers a potential solution,^12–14^ yet historically computer tools have been limited by simplistic scoring rubrics and high development costs.^15, 16^ Artificial intelligence (AI) large language models (LLMs) such as OpenAI GPT lower these barriers,^17–21^ but the accuracy, reliability, and cost of these tools remains incompletely understood. Moreover, best practices such as the specific wording of LLM instructions (“prompts”) lack solid evidence.^18, 22–24^ Educators, researchers, and computer scientists would benefit from evidence regarding LLM scoring of narrative assessments, and how outcomes vary for different prompts (i.e., prompt-writing best practices). To address these gaps, the present study evaluated various LLM-powered approaches for scoring medical student reflective essays.

Computers have been used to score essays since 1966,^12, 13^ and the field of Automated Essay Scoring (AES) has developed a robust literature with numerous commercial and bespoke tools.^15, 25, 26^ Automated scoring alleviates human rater burden^20^ and improves student performance on writing tasks.^27^ However, traditional AES is limited by demands of technical expertise, monetary investment, and model-specific development.^13, 15, 26^ Most AES systems are content-generic, and rely on surrogate measures of quality such as grammar and mechanics. Systems that incorporate higher-order features such as content and reasoning have historically required resource-intensive, content-specific scoring algorithms.^15, 26^ Machine learning can help by “training” a novel algorithm, but typically requires in-depth technical expertise and numerous manually classified (labeled or annotated) examples. Thus, as recently as 2019 one reviewer concluded “Content is largely beyond the reach of state-of-the-art essay scoring engines.”^15^

Contemporary AI approaches, including LLMs, may bypass these obstacles.^17, 18^ First, LLMs largely obviate the need for technical skill in machine learning, substituting instead the need to carefully craft an effective prompt. Second, LLMs’ ability to interpret narrative text may reduce the number of required human-rated examples.^22^ Moreover, LLMs excel in generating narrative responses, which could be useful in providing detailed written feedback to learners.^24^ The wide availability of LLMs also extends access to educators with limited funds or technical expertise.

In medicine, computer-based scoring of narrative assessments has been used in a wide range of educational and clinical contexts.^16, 17^ However, as with AES, AI-based ratings have historically required technical expertise and numerous human-rated exemplars (for example^28–31^). Recent studies using OpenAI GPT and other LLMs found generally favorable results (e.g., agreement with human scores) with little or no pre-training when scoring clinician dialogs with virtual patients;^19^ medical student essays,^20, 32^ post-encounter notes,^21, 33^ and conversations with standardized patients;^34^ and dental student care plans.^35^

With proof-of-concept now demonstrated, a vital next step is to investigate LLM prompt engineering strategies. Identifying generalizable best practices for effective (accurate, reproducible) and efficient (low cost) scoring is essential for widespread adoption of LLM-powered assessment. Moreover, best-practice prompt templates would enable educators without technical expertise to use these tools. In medical education, few studies have compared prompt variations: 1 compared the use of exemplars (few-shot or in-context learning) vs none (zero-shot learning),^32^ and another compared analysis of full text conversations vs a text extract^34^). A third study reported resource requirements (money, time) for different LLM models.^19^ Looking beyond medicine, studies have explored providing exemplars vs none;^18, 36–38^ using “chain-of-thought” prompts;^22, 23, 39^ using the LLM to auto-generate or modify exemplars or chain-of-thought rationales;^39–41^ and asking the LLM to provide narrative feedback before the numeric score.^18^ However, studies are few, results are inconsistent, and many were done using now-archaic LLMs.

Thus, *it remains unknown* how to structure prompts to optimize performance and cost. Narrowing this gap would illuminate best practices for LLM-powered automated scoring of narrative assessments. The purpose of this study was to evaluate GPT for scoring medical student reflective essays, and compare various prompting techniques and GPT models in terms of accuracy, reproducibility, and cost.

## Methods

We used OpenAI GPT to score 51 medical student reflection essays (15 real, 36 fabricated) in April-May 2025. We systematically compared various scoring conditions (LLM prompts and models) using outcomes of accuracy (compared with human raters), reproducibility (across replications using the same condition), and cost (money, time). See Table 1 for specific prompt-model conditions, hypotheses, and verbatim prompts. No human subjects were enrolled in this study.

**Table 1.**
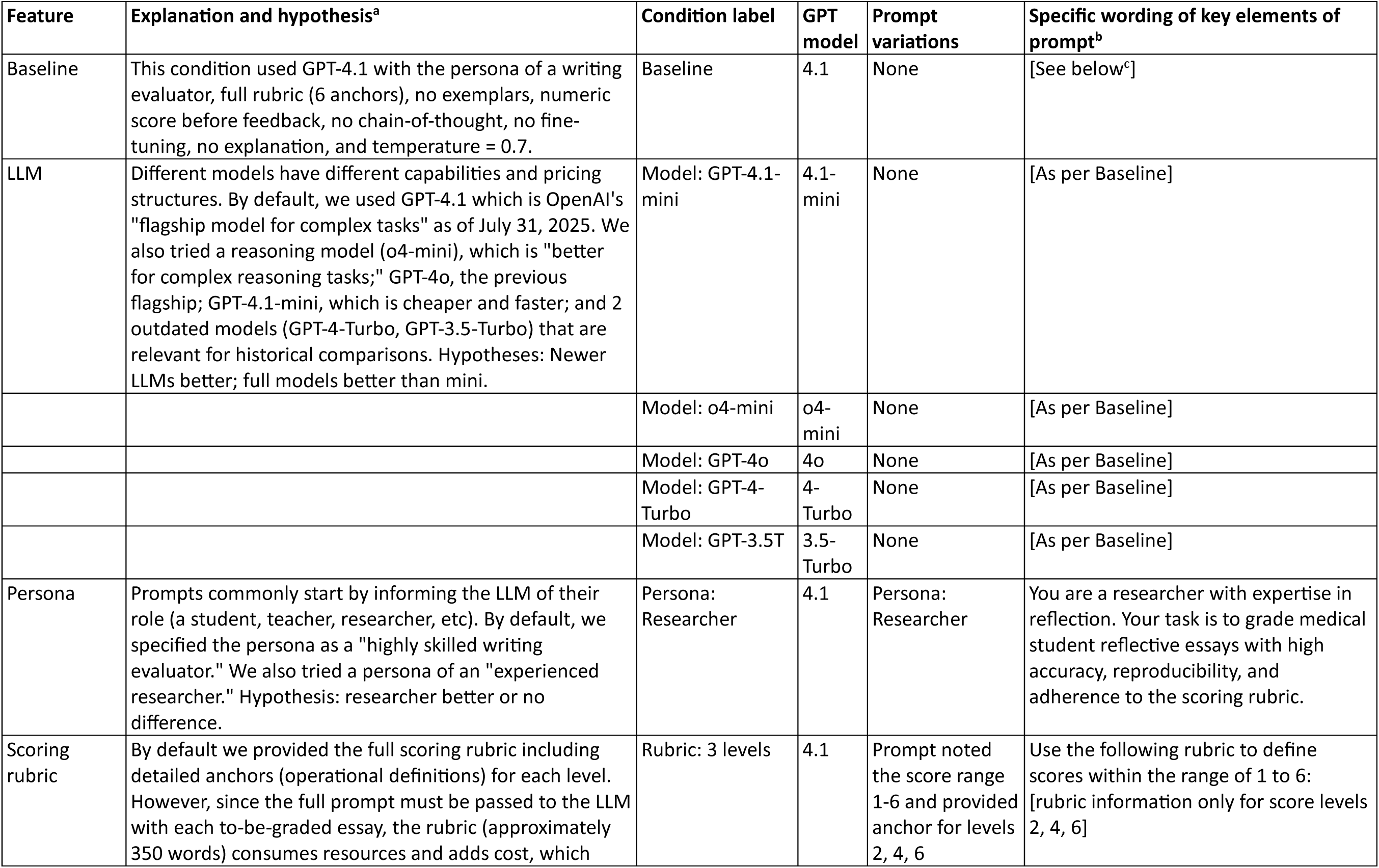
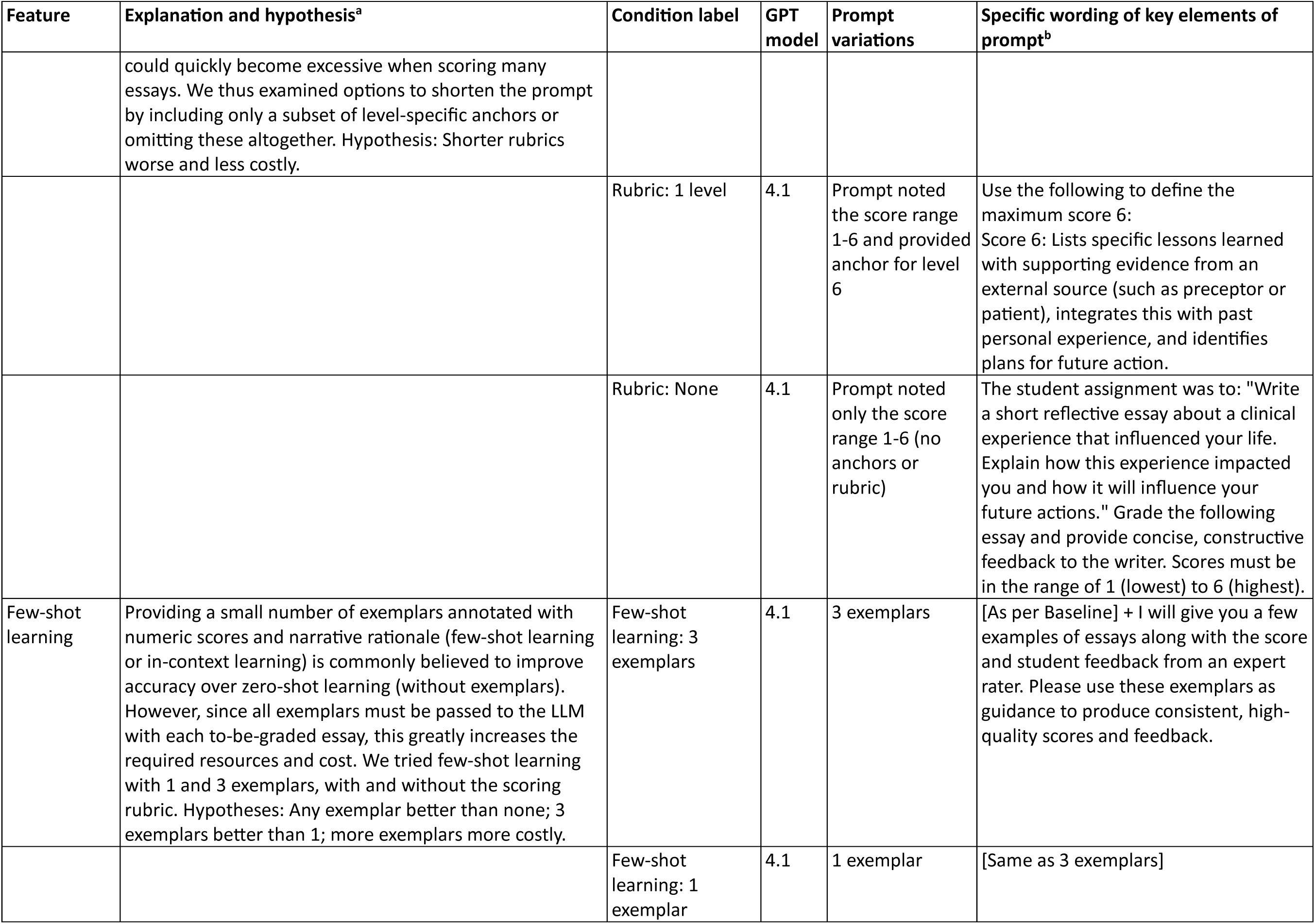
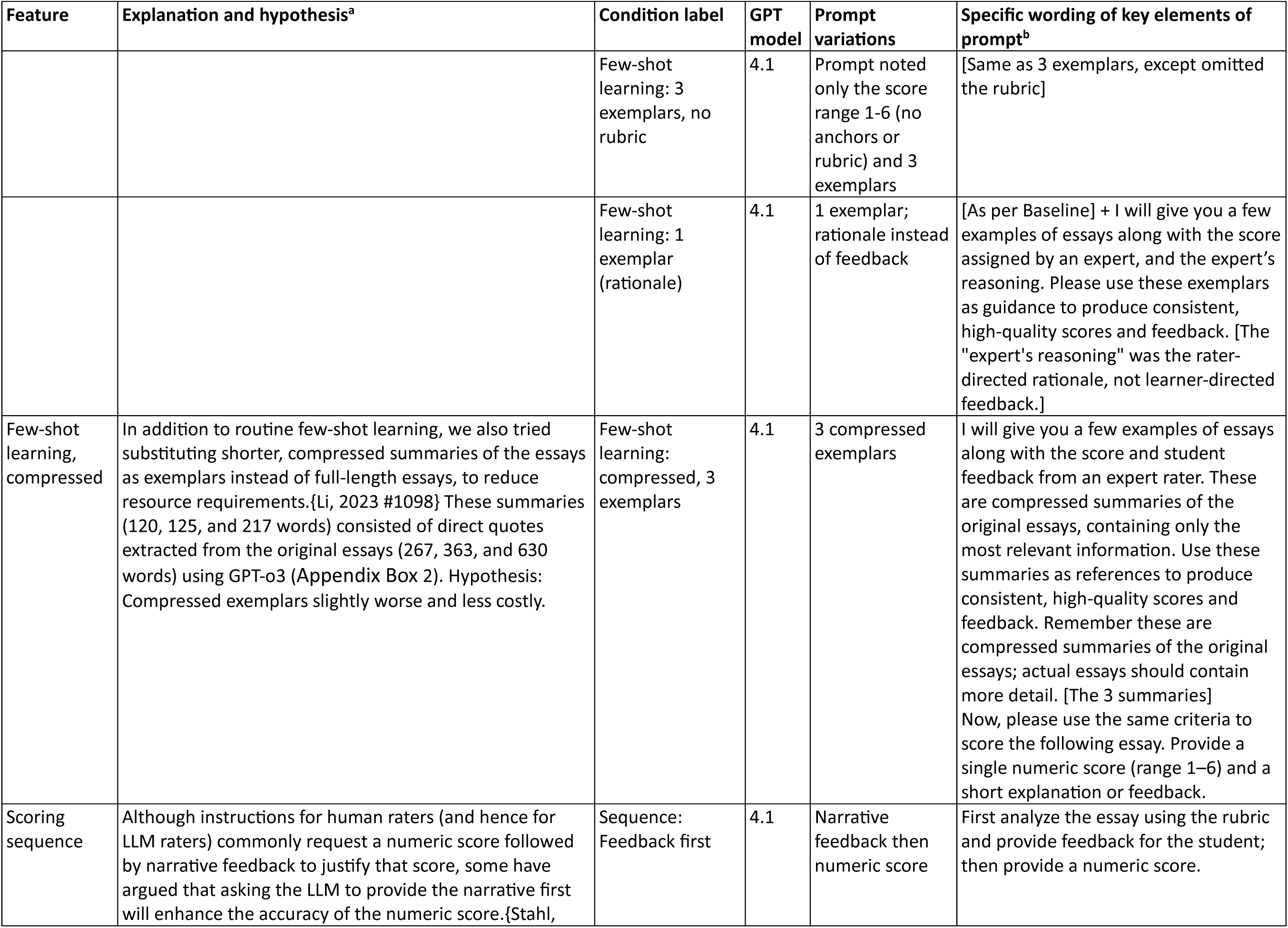
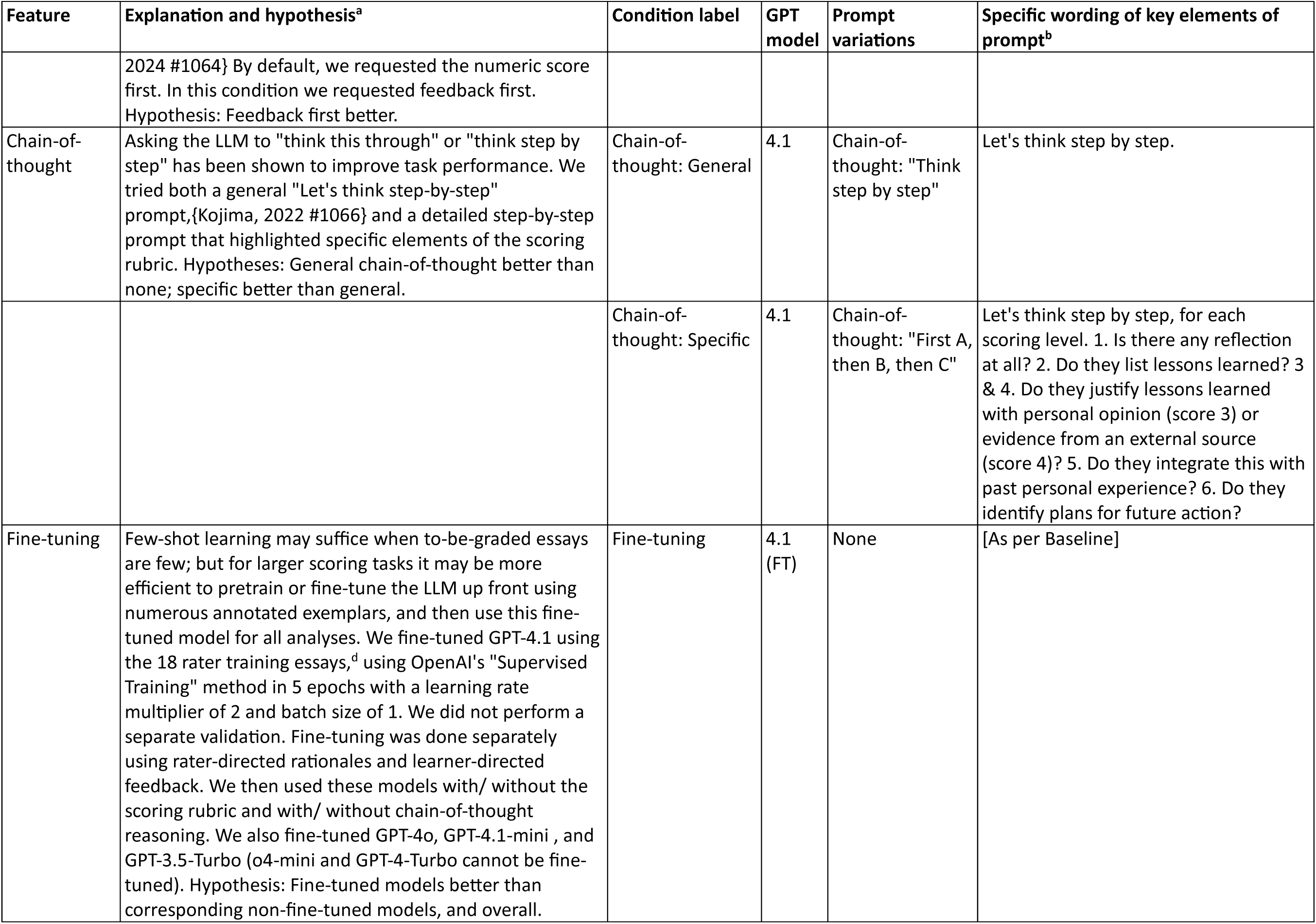
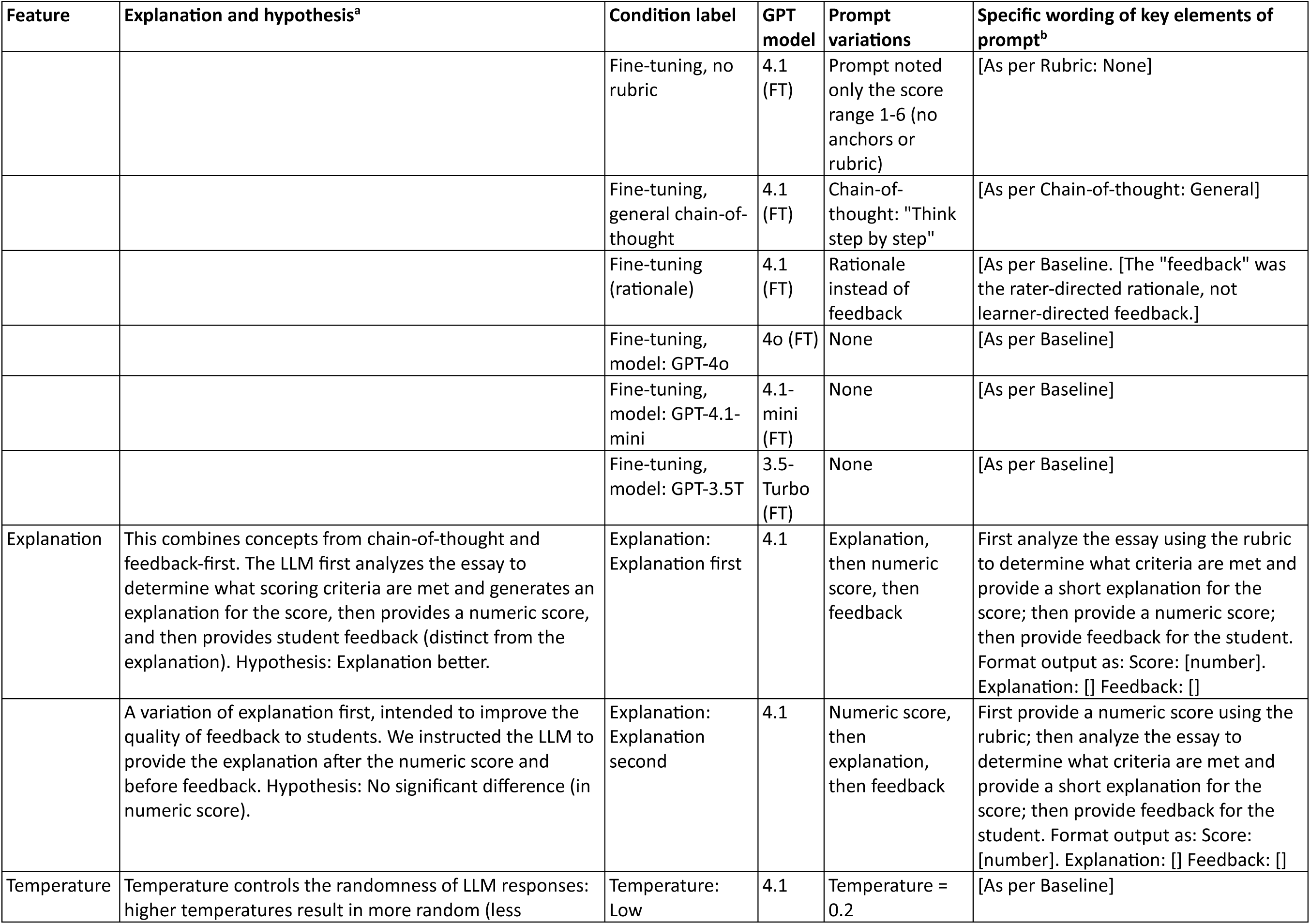
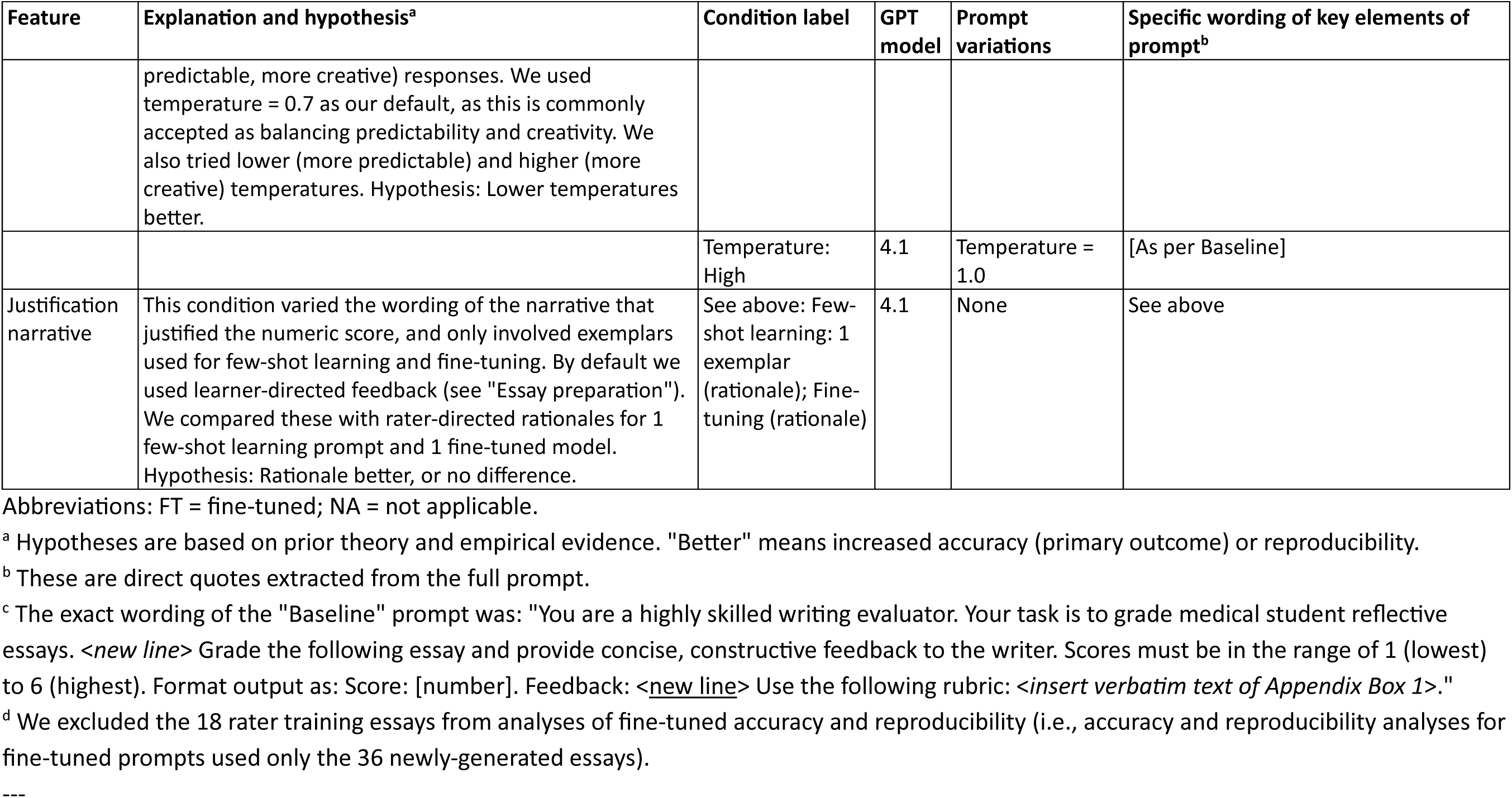
Definitions and wording of prompt-model conditions used to score reflection essays using large language models.

| Feature | Explanation and hypothesis<sup>a</sup> | Condition label | GPT model | Prompt variations | Specific wording of key elements of prompt<sup>b</sup> |
| --- | --- | --- | --- | --- | --- |
| Baseline | This condition used GPT-4.1 with the persona of a writing evaluator, full rubric (6 anchors), no exemplars, numeric score before feedback, no chain-of-thought, no fine-tuning, no explanation, and temperature = 0.7. | Baseline | 4.1 | None | [See below<sup>c</sup>] |
| LLM | Different models have different capabilities and pricing structures. By default, we used GPT-4.1 which is OpenAI's "flagship model for complex tasks" as of July 31, 2025. We also tried a reasoning model (o4-mini), which is "better for complex reasoning tasks;" GPT-4o, the previous flagship; GPT-4.1-mini, which is cheaper and faster; and 2 outdated models (GPT-4-Turbo, GPT-3.5-Turbo) that are relevant for historical comparisons. Hypotheses: Newer LLMs better; full models better than mini. | Model: GPT-4.1-mini | 4.1-mini | None | [As per Baseline] |
|  |  | Model: o4-mini | o4-mini | None | [As per Baseline] |
|  |  | Model: GPT-4o | 4o | None | [As per Baseline] |
|  |  | Model: GPT-4-Turbo | 4-Turbo | None | [As per Baseline] |
|  |  | Model: GPT-3.5T | 3.5-Turbo | None | [As per Baseline] |
| Persona | Prompts commonly start by informing the LLM of their role (a student, teacher, researcher, etc). By default, we specified the persona as a "highly skilled writing evaluator." We also tried a persona of an "experienced researcher." Hypothesis: researcher better or no difference. | Persona: Researcher | 4.1 | Persona: Researcher | You are a researcher with expertise in reflection. Your task is to grade medical student reflective essays with high accuracy, reproducibility, and adherence to the scoring rubric. |
| Scoring rubric | By default we provided the full scoring rubric including detailed anchors (operational definitions) for each level. However, since the full prompt must be passed to the LLM with each to-be-graded essay, the rubric (approximately 350 words) consumes resources and adds cost, which could quickly become excessive when scoring many essays. We thus examined options to shorten the prompt by including only a subset of level-specific anchors or omitting these altogether. Hypothesis: Shorter rubrics worse and less costly. | Rubric: 3 levels | 4.1 | Prompt noted the score range 1-6 and provided anchor for levels 2, 4, 6 | Use the following rubric to define scores within the range of 1 to 6: [rubric information only for score levels 2, 4, 6] |
|  |  | Rubric: 1 level | 4.1 | Prompt noted the score range 1-6 and provided anchor for level 6 | Use the following to define the maximum score 6:<br>Score 6: Lists specific lessons learned with supporting evidence from an external source (such as preceptor or patient), integrates this with past personal experience, and identifies plans for future action. |
|  |  | Rubric: None | 4.1 | Prompt noted only the score range 1-6 (no anchors or rubric) | The student assignment was to: "Write a short reflective essay about a clinical experience that influenced your life. Explain how this experience impacted you and how it will influence your future actions." Grade the following essay and provide concise, constructive feedback to the writer. Scores must be in the range of 1 (lowest) to 6 (highest). |
| Few-shot learning | Providing a small number of exemplars annotated with numeric scores and narrative rationale (few-shot learning or in-context learning) is commonly believed to improve accuracy over zero-shot learning (without exemplars). However, since all exemplars must be passed to the LLM with each to-be-graded essay, this greatly increases the required resources and cost. We tried few-shot learning with 1 and 3 exemplars, with and without the scoring rubric. Hypotheses: Any exemplar better than none; 3 exemplars better than 1; more exemplars more costly. | Few-shot learning: 3 exemplars | 4.1 | 3 exemplars | [As per Baseline] + I will give you a few examples of essays along with the score and student feedback from an expert rater. Please use these exemplars as guidance to produce consistent, high-quality scores and feedback. |
|  |  | Few-shot learning: 1 exemplar | 4.1 | 1 exemplar | [Same as 3 exemplars] |
|  |  | Few-shot learning: 3 exemplars, no rubric | 4.1 | Prompt noted only the score range 1-6 (no anchors or rubric) and 3 exemplars | [Same as 3 exemplars, except omitted the rubric] |
|  |  | Few-shot learning: 1 exemplar (rationale) | 4.1 | 1 exemplar; rationale instead of feedback | [As per Baseline] + I will give you a few examples of essays along with the score assigned by an expert, and the expert's reasoning. Please use these exemplars as guidance to produce consistent, high-quality scores and feedback. [The "expert's reasoning" was the rater-directed rationale, not learner-directed feedback.] |
| Few-shot learning, compressed | In addition to routine few-shot learning, we also tried substituting shorter, compressed summaries of the essays as exemplars instead of full-length essays, to reduce resource requirements.{Li, 2023 #1098} These summaries (120, 125, and 217 words) consisted of direct quotes extracted from the original essays (267, 363, and 630 words) using GPT-o3 (Appendix Box 2). Hypothesis: Compressed exemplars slightly worse and less costly. | Few-shot learning: compressed, 3 exemplars | 4.1 | 3 compressed exemplars | I will give you a few examples of essays along with the score and student feedback from an expert rater. These are compressed summaries of the original essays, containing only the most relevant information. Use these summaries as references to produce consistent, high-quality scores and feedback. Remember these are compressed summaries of the original essays; actual essays should contain more detail. [The 3 summaries] Now, please use the same criteria to score the following essay. Provide a single numeric score (range 1–6) and a short explanation or feedback. |
| Scoring sequence | Although instructions for human raters (and hence for LLM raters) commonly request a numeric score followed by narrative feedback to justify that score, some have argued that asking the LLM to provide the narrative first will enhance the accuracy of the numeric score.{Stahl, | Sequence: Feedback first | 4.1 | Narrative feedback then numeric score | First analyze the essay using the rubric and provide feedback for the student; then provide a numeric score. |
|  | 2024 #1064} By default, we requested the numeric score first. In this condition we requested feedback first.<br>Hypothesis: Feedback first better. |  |  |  |  |
| Chain-of-thought | Asking the LLM to "think this through" or "think step by step" has been shown to improve task performance. We tried both a general "Let's think step-by-step" prompt,{Kojima, 2022 #1066} and a detailed step-by-step prompt that highlighted specific elements of the scoring rubric. Hypotheses: General chain-of-thought better than none; specific better than general. | Chain-of-thought: General | 4.1 | Chain-of-thought: "Think step by step" | Let's think step by step. |
|  |  | Chain-of-thought: Specific | 4.1 | Chain-of-thought: "First A, then B, then C" | Let's think step by step, for each scoring level. 1. Is there any reflection at all? 2. Do they list lessons learned? 3 & 4. Do they justify lessons learned with personal opinion (score 3) or evidence from an external source (score 4)? 5. Do they integrate this with past personal experience? 6. Do they identify plans for future action? |
| Fine-tuning | Few-shot learning may suffice when to-be-graded essays are few; but for larger scoring tasks it may be more efficient to pretrain or fine-tune the LLM up front using numerous annotated exemplars, and then use this fine-tuned model for all analyses. We fine-tuned GPT-4.1 using the 18 rater training essays,<sup>d</sup> using OpenAI's "Supervised Training" method in 5 epochs with a learning rate multiplier of 2 and batch size of 1. We did not perform a separate validation. Fine-tuning was done separately using rater-directed rationales and learner-directed feedback. We then used these models with/ without the scoring rubric and with/ without chain-of-thought reasoning. We also fine-tuned GPT-4o, GPT-4.1-mini , and GPT-3.5-Turbo (o4-mini and GPT-4-Turbo cannot be fine-tuned). Hypothesis: Fine-tuned models better than corresponding non-fine-tuned models, and overall. | Fine-tuning | 4.1 (FT) | None | [As per Baseline] |
|  |  | Fine-tuning, no rubric | 4.1 (FT) | Prompt noted only the score range 1-6 (no anchors or rubric) | [As per Rubric: None] |
|  |  | Fine-tuning, general chain-of-thought | 4.1 (FT) | Chain-of-thought: "Think step by step" | [As per Chain-of-thought: General] |
|  |  | Fine-tuning (rationale) | 4.1 (FT) | Rationale instead of feedback | [As per Baseline. [The "feedback" was the rater-directed rationale, not learner-directed feedback.]] |
|  |  | Fine-tuning, model: GPT-4o | 4o (FT) | None | [As per Baseline] |
|  |  | Fine-tuning, model: GPT-4.1-mini | 4.1-mini (FT) | None | [As per Baseline] |
|  |  | Fine-tuning, model: GPT-3.5T | 3.5-Turbo (FT) | None | [As per Baseline] |
| Explanation | This combines concepts from chain-of-thought and feedback-first. The LLM first analyzes the essay to determine what scoring criteria are met and generates an explanation for the score, then provides a numeric score, and then provides student feedback (distinct from the explanation). Hypothesis: Explanation better. | Explanation: Explanation first | 4.1 | Explanation, then numeric score, then feedback | First analyze the essay using the rubric to determine what criteria are met and provide a short explanation for the score; then provide a numeric score; then provide feedback for the student. Format output as: Score: [number]. Explanation: [] Feedback: [] |
|  | A variation of explanation first, intended to improve the quality of feedback to students. We instructed the LLM to provide the explanation after the numeric score and before feedback. Hypothesis: No significant difference (in numeric score). | Explanation: Explanation second | 4.1 | Numeric score, then explanation, then feedback | First provide a numeric score using the rubric; then analyze the essay to determine what criteria are met and provide a short explanation for the score; then provide feedback for the student. Format output as: Score: [number]. Explanation: [] Feedback: [] |
| Temperature | Temperature controls the randomness of LLM responses: higher temperatures result in more random (less predictable, more creative) responses. We used temperature = 0.7 as our default, as this is commonly accepted as balancing predictability and creativity. We also tried lower (more predictable) and higher (more creative) temperatures. Hypothesis: Lower temperatures better. | Temperature: Low | 4.1 | Temperature = 0.2 | [As per Baseline] |
|  |  | Temperature: High | 4.1 | Temperature = 1.0 | [As per Baseline] |
| Justification narrative | This condition varied the wording of the narrative that justified the numeric score, and only involved exemplars used for few-shot learning and fine-tuning. By default we used learner-directed feedback (see "Essay preparation"). We compared these with rater-directed rationales for 1 few-shot learning prompt and 1 fine-tuned model. Hypothesis: Rationale better, or no difference. | See above: Few-shot learning: 1 exemplar (rationale); Fine-tuning (rationale) | 4.1 | None | See above |

Abbreviations: FT = fine-tuned; NA = not applicable.
<sup>a</sup> Hypotheses are based on prior theory and empirical evidence. "Better" means increased accuracy (primary outcome) or reproducibility.
<sup>b</sup> These are direct quotes extracted from the full prompt.
<sup>c</sup> The exact wording of the "Baseline" prompt was: "You are a highly skilled writing evaluator. Your task is to grade medical student reflective essays. <new line> Grade the following essay and provide concise, constructive feedback to the writer. Scores must be in the range of 1 (lowest) to 6 (highest). Format output as: Score: [number]. Feedback: <new line> Use the following rubric: <insert verbatim text of Appendix Box 1>."
<sup>d</sup> We excluded the 18 rater training essays from analyses of fine-tuned accuracy and reproducibility (i.e., accuracy and reproducibility analyses for fine-tuned prompts used only the 36 newly-generated essays).

### Scoring rubric and rater training

We used the “Reflective Ability Rubric,”^42^ an open-source resource for scoring medical student reflection essays. Validity of scores generated using this rubric (Appendix Box 1) is supported by evidence of content, internal structure, and relations with other variables. The rubric was grounded in a framework in which high-quality reflection includes specific lessons learned, external evidence to justify these lessons as meaningful, integration with other previous experiences, and plans for future actions. In the original validation, inter-rater reliability was high, and higher scores were associated with later stages of training and higher assessments of professionalism and communication skills.

The published resource includes 18 rater training essays (3 essays for each scoring level) and a brief rationale for each. Investigator TAL trained using all 18 essays. Investigator DAC trained using only 6 essays, because he planned to later confirm the published scores and wanted to minimize bias.

### Essay preparation

The 51 essays used in this study (length 104-868 words) were previously published or fabricated for research purposes. The writing task asked a clinical-year medical student to “Write a short reflective essay about a clinical experience that influenced your life. Explain how this experience impacted you and how it will influence your future actions.”

We used the 18 rater training essays included with the published rubric.^42^ We confirmed with the author that these were real student essays (personal communication, P.S. O’Sullivan, 21 November 2025). We selected 3 essays as exemplars (at scoring levels 2, 4, and 6) for few-shot learning, and used the remaining 15 for testing. To confirm the accuracy of the published scores, 3 weeks after training author DAC scored all 18 essays while blinded to the original scores, with 100% agreement. We then used these scores as the reference standard.

The other 36 essays were generated by a different LLM (Gemini-2.0-flash) on April 18, 2025. For each of the 18 rater training (real) essays we asked Gemini to create 2 novel essays representing new clinical scenarios while preserving the original level of reflection. The full investigator-LLM prompting dialog is shown in Appendix Table A1. The prompt requested (in part): “Generate a new set of reflective essays that are similar in their writing quality to the original 18. … Vary the medical specialty,… patient,… and clinical condition. … I want the depth of reflection to match each example essay.” The scoring rubric including operational definitions was then uploaded, along with the original essays. We noted that the generated level 1 and 2 essays were incongruously good, whereas level 5 essays lacked differentiation from level 4. We thus directed Gemini to rewrite these essays, using prompts that stated, in part, the following. Level 1: “Rewrite it to make it lower quality … make it not reflective.” Level 2: “Retain a little bit of reflection … but it will be very poor reflection, very vague.” Level 5: “[The essays] lack external evidence … remove any reference to future plans or implications … add a lesson learned (without specifically mentioning the future).” The generated essays were independently scored by 2 investigators (authors DAC, TAL) blinded to the intended score. These raters agreed on scores for 34 of 36 newly-generated essays (raw agreement 94%; kappa 0.93), and then reconciled differences to create reference-standard scores.

The 18 rater training essays were longer (mean [SD] 432 [150] words) than the 36 Gemini-generated essays (239 [89]). Higher-scored essays (422 [201] words for level 6) also tended to be longer than lower-scored essays (256 [81] for level 1), although there was substantial variability in this pattern; see details in Appendix Table A2.

### LLM scoring

We asked GPT to “grade the following essay and provide concise, constructive feedback to the writer,” using several different scoring conditions for each essay (Table 1). We replicated each condition 6 times. We accessed GPT through the OpenAI application programming interface (API) using a short Python script. We recorded the time and number of tokens required. We calculated cost in US dollars based on OpenAI per-token pricing (see Appendix Table A3).

### Variation in scoring conditions

As guided by theories and best practices for prompt engineering, we systematically planned 29 prompt-model conditions by varying the persona, scoring rubric, GPT model, few-shot learning (use of exemplars, use of compressed exemplars), fine-tuning, scoring sequence (numeric vs feedback first), chain-of-thought reasoning, explanation, temperature, and feedback/ rationale. See Table 1 for details. We used GPT-o3 to generate the 3 compressed training essays (full prompt shown in Appendix Box 2).

We initially used the published rationales as exemplars and for fine-tuning, but this led GPT to offer “feedback” phrased as rater-directed justifications for scoring rather than learner-directed feedback. To address this, author DAC revised each training essay rationale into learner-directed feedback. We used feedback as the default for all exemplar prompts and fine-tuning, retaining rater-directed rationales for 2 conditions (1 each for exemplars and fine-tuning).

### Data analysis

The outcome of primary interest was accuracy – the degree to which the LLM scores agreed with reference-standard human scores. We measured accuracy using single-score intraclass correlation coefficient (ICC)-like measures (comparable to kappa), calculated from variance components reflecting within-essay and between-rater (human vs LLM) variability. We additionally measured accuracy (for formal statistical analyses and between-condition comparisons) as the absolute value of the difference between LLM-assigned scores and corresponding human scores, and averaged across replicate assessments to compute mean absolute differences (MADs). We used analysis of variance approaches to contrast MADs among different prompt-model conditions while adjusting for differences among the essays. After performing a global test of significance among all conditions, we estimated differences and 95% confidence intervals (CIs) for specific comparisons of interest. We maintained false discovery (type I error) at a per-comparison level of 0.05 using the Benjamini-Hochberg procedure.^43^ We evaluated the reproducibility of LLM scores across replications using single-score ICCs. We interpreted ICCs using thresholds proposed by Landis and Koch^44^ which include 0.61-0.8 substantial and >0.8 almost perfect. All analyses were conducted using SAS (Cary, NC).

## Results

### Score distributions

LLM-generated scores were, on average, slightly higher than human reference standard scores for all 51 essays (mean [SD], 3.74 [1.58] across 306 “baseline” condition scores; vs 3.53 [1.68] across 51 human scores), and for the 36 Gemini-generated essays (3.73 [1.77], N=216; vs 3.58 [1.66], N=36). See Appendix Table A2 for counts for each score level.

### Resource requirements

We scored 51 essays using 29 different conditions (different prompts or models), with 6 replications each (i.e., N=306 data points for each condition except fine-tuned models, for which N=216 [36 essays]). Across all conditions, it took mean (SD) 3.73 (3.12) seconds and 902 (169) tokens for GPT to score 1 essay. Average time for specific conditions (see Table 2) ranged from 1.98 sec (for GPT-4.1-mini) to 6.76 sec (for explanation second). Likewise, the total number of tokens varied across conditions from 545 (for no rubric, fine-tuned) to 2503 (for 3 exemplars). A sensitivity analysis limited to the 36 essays not used for fine-tuning revealed very similar results (overall slightly faster and cheaper, but essentially the same rank order of conditions; data not shown).

**Table 2.** Resources required to score reflection essays using large language models for various prompt-model conditions^a^.

| Condition | GPT model | Cost (USD per 100 essays), N=10,000<sup>b</sup> | Cost (USD per 100 essays), N=100<sup>b</sup> | Total tokens, mean (SD) | Prompt tokens, mean (SD) | Completion tokens, mean (SD) | Time in sec, mean (SD) |
| --- | --- | --- | --- | --- | --- | --- | --- |
| Baseline + FT model variation | 4.1-mini (FT) | 0.04 | 0.40 | 635 (162) | 558 (152) | 77 (36) | 1.98 (0.66) |
| Baseline + model variation | 4.1-mini | 0.04 | 0.04 | 672 (161) | 558 (152) | 113 (16) | 2.62 (0.80) |
| Baseline + model variation | 3.5-Turbo | 0.04 | 0.04 | 670 (165) | 564 (154) | 106 (22) | 2.20 (0.55) |
| Baseline + FT model variation | 3.5-Turbo (FT) | 0.04 | 0.63 | 634 (160) | 564 (154) | 71 (30) | 2.94 (7.93) |
| Rubric: None + FT | 4.1 (FT) | 0.18 | 2.00 | 545 (201) | 449 (152) | 95 (114) | 2.72 (1.78) |
| Baseline (rationale) + FT model variation | 4.1 (FT) | 0.19 | 2.00 | 632 (159) | 558 (152) | 73 (31) | 2.26 (0.61) |
| Chain-of-thought: General + FT | 4.1 (FT) | 0.20 | 2.01 | 648 (158) | 564 (152) | 84 (31) | 2.53 (0.83) |
| Baseline + FT model variation | 4.1 (FT) | 0.20 | 2.01 | 646 (158) | 558 (152) | 88 (32) | 2.67 (0.84) |
| Rubric: 3 levels | 4.1 | 0.20 | 0.20 | 630 (167) | 500 (152) | 129 (21) | 3.47 (1.42) |
| Rubric: None | 4.1 | 0.21 | 0.21 | 597 (170) | 449 (152) | 147 (24) | 4.18 (1.47) |
| Rubric: 1 level | 4.1 | 0.21 | 0.21 | 608 (168) | 460 (152) | 148 (23) | 4.14 (1.64) |
| Temperature: Low | 4.1 | 0.21 | 0.21 | 683 (167) | 558 (152) | 124 (20) | 3.75 (1.17) |
| Baseline | 4.1 | 0.21 | 0.21 | 684 (167) | 558 (152) | 125 (21) | 3.58 (1.14) |
| Chain-of-thought: General | 4.1 | 0.21 | 0.21 | 689 (169) | 564 (152) | 125 (23) | 4.64 (2.09) |
| Persona: Researcher | 4.1 | 0.21 | 0.21 | 697 (167) | 572 (152) | 124 (21) | 3.30 (0.99) |
| Temperature: High | 4.1 | 0.21 | 0.21 | 687 (169) | 558 (152) | 129 (23) | 3.87 (1.28) |
| Sequence: Feedback first | 4.1 | 0.23 | 0.23 | 724 (170) | 578 (152) | 145 (24) | 4.12 (5.07) |
| Chain-of-thought: Specific | 4.1 | 0.23 | 0.23 | 773 (168) | 641 (152) | 132 (24) | 5.19 (5.52) |
| Baseline + FT model variation | 4o (FT) | 0.23 | 2.05 | 635 (158) | 558 (152) | 77 (24) | 3.13 (5.98) |
| Baseline + model variation | 4o | 0.26 | 0.26 | 684 (161) | 558 (152) | 125 (18) | 3.52 (0.79) |
| Explanation: Explanation second | 4.1 | 0.27 | 0.27 | 788 (174) | 597 (152) | 191 (29) | 6.76 (34.55) |
| Explanation: Explanation first | 4.1 | 0.27 | 0.27 | 789 (175) | 597 (152) | 191 (32) | 4.81 (1.80) |
| Baseline + model variation | o4-mini | 0.30 | 0.30 | 1,092 (254) | 557 (152) | 535 (167) | 6.60 (2.25) |
| Few-shot learning: 1 exemplar (rationale) | 4.1 | 0.38 | 0.38 | 1,527 (167) | 1,409 (152) | 118 (22) | 3.84 (2.03) |
| Few-shot learning: 1 exemplar | 4.1 | 0.38 | 0.38 | 1,557 (166) | 1,436 (152) | 121 (21) | 3.01 (0.93) |
| Few-shot learning: compressed, 3 exemplars | 4.1 | 0.40 | 0.40 | 1,647 (164) | 1,521 (152) | 126 (17) | 3.50 (1.27) |
| Few-shot learning: 3 exemplars, no rubric | 4.1 | 0.55 | 0.55 | 2,368 (163) | 2,241 (152) | 126 (16) | 4.26 (1.66) |
| Few-shot learning: 3 exemplars | 4.1 | 0.57 | 0.57 | 2,503 (164) | 2,382 (152) | 120 (17) | 4.01 (2.39) |
| Baseline + model variation | 4-Turbo | 0.98 | 0.98 | 703 (164) | 564 (154) | 139 (24) | 4.43 (1.12) |

<sup>a</sup> Time and tokens are mean for 1 essay. Data represent the mean (SD) across 6 replications (N=306 data points [51 essays], except for fine-tuned models N=216 data points [36 essays]). Analyses limited to the 36 essays scored in all conditions revealed very similar results. See Table 1 for details on conditions.
<sup>b</sup> We calculated cost using pricing as per OpenAI, May 1, 2025 (see Appendix Table A3 for details). We report cost for 2 situations: scoring 10,000 essays and scoring 100 essays. To facilitate comparison, we report the cost for both situations as standardized to 100 essays. Fine-tuned models include the one-time cost of fine-tuning, which when amortized across 10,000 essays results in a lower per-essay cost.

The cost to score 100 essays for the baseline condition (GPT-4.1 with full rubric and no exemplars) was USD $0.21, and ranged from $0.04 (for GPT-4.1-mini) to $2.05 (for fine-tuned GPT-4o). The 5 most expensive conditions for the 100-essay task were all fine-tuned models (each ≥$2.00). However, the cost of fine-tuning is a one-time up-front expense that ranged $0.37 for GPT-4.1-mini to $1.83 for GPT-4.1 and GPT-4o. When amortized over 10,000 essays the per-essay cost was $3.83 per 10,000 ($0.04 per 100) for fine-tuned GPT-4.1-mini, and $0.20 per 100 for fine-tuned GPT-4.1.

### Accuracy

LLM accuracy, as measured using ICC, was “almost perfect” (>0.80) for 28/29 conditions (97%), and >0.90 for 19 (66%); see Figure 1. Accuracy ICCs ranged from 0.97 (for fine-tuned with rationale) to 0.78 (for no rubric). The most accurate conditions were fine-tuned models (4 of the top 7) and prompts with exemplars (i.e., few-shot learning; 5 of the top 9). However, a fine-tuned model (using the much older GPT-3.5-Turbo) was also the third-least accurate condition.

**Figure 1.**
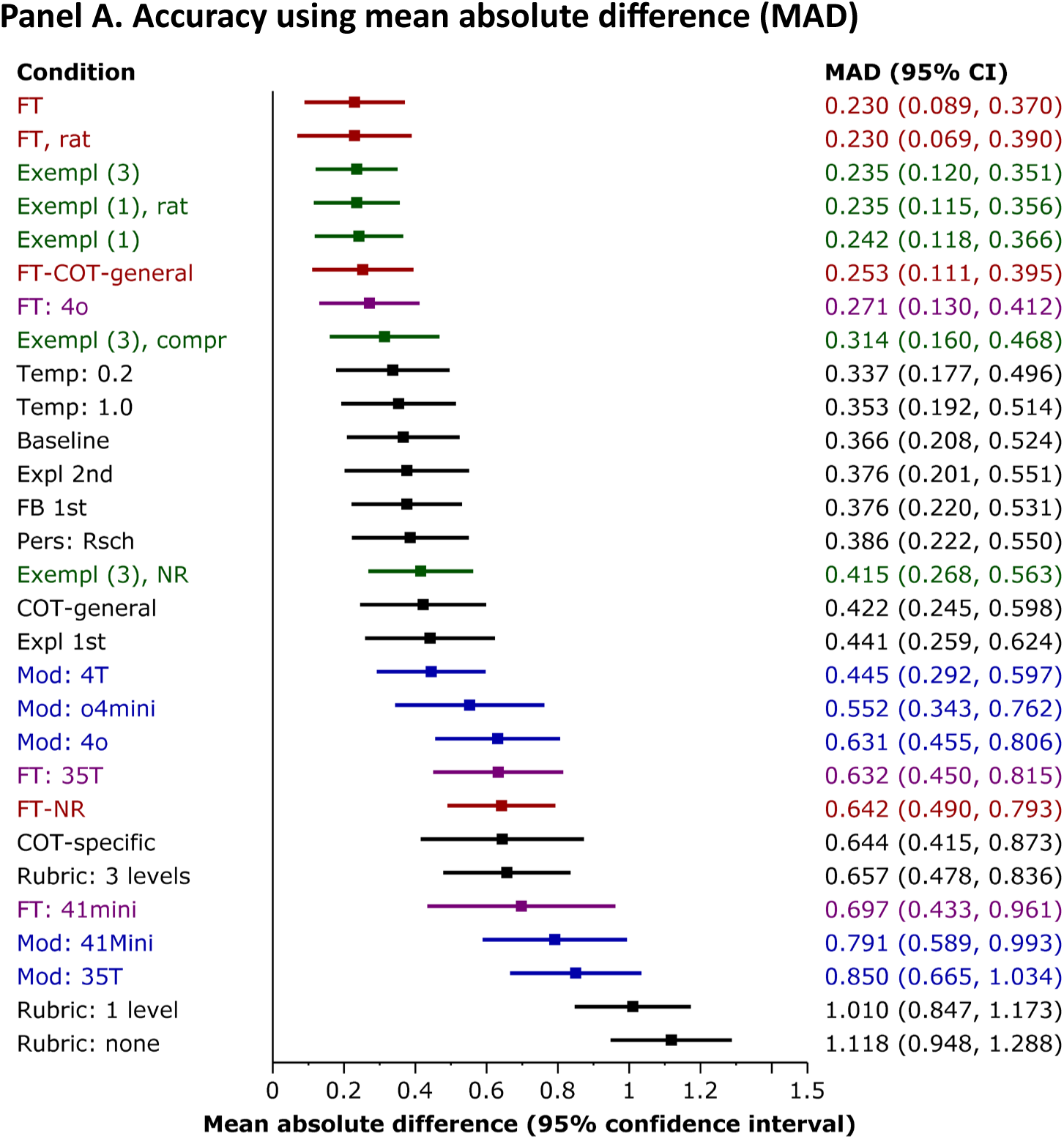
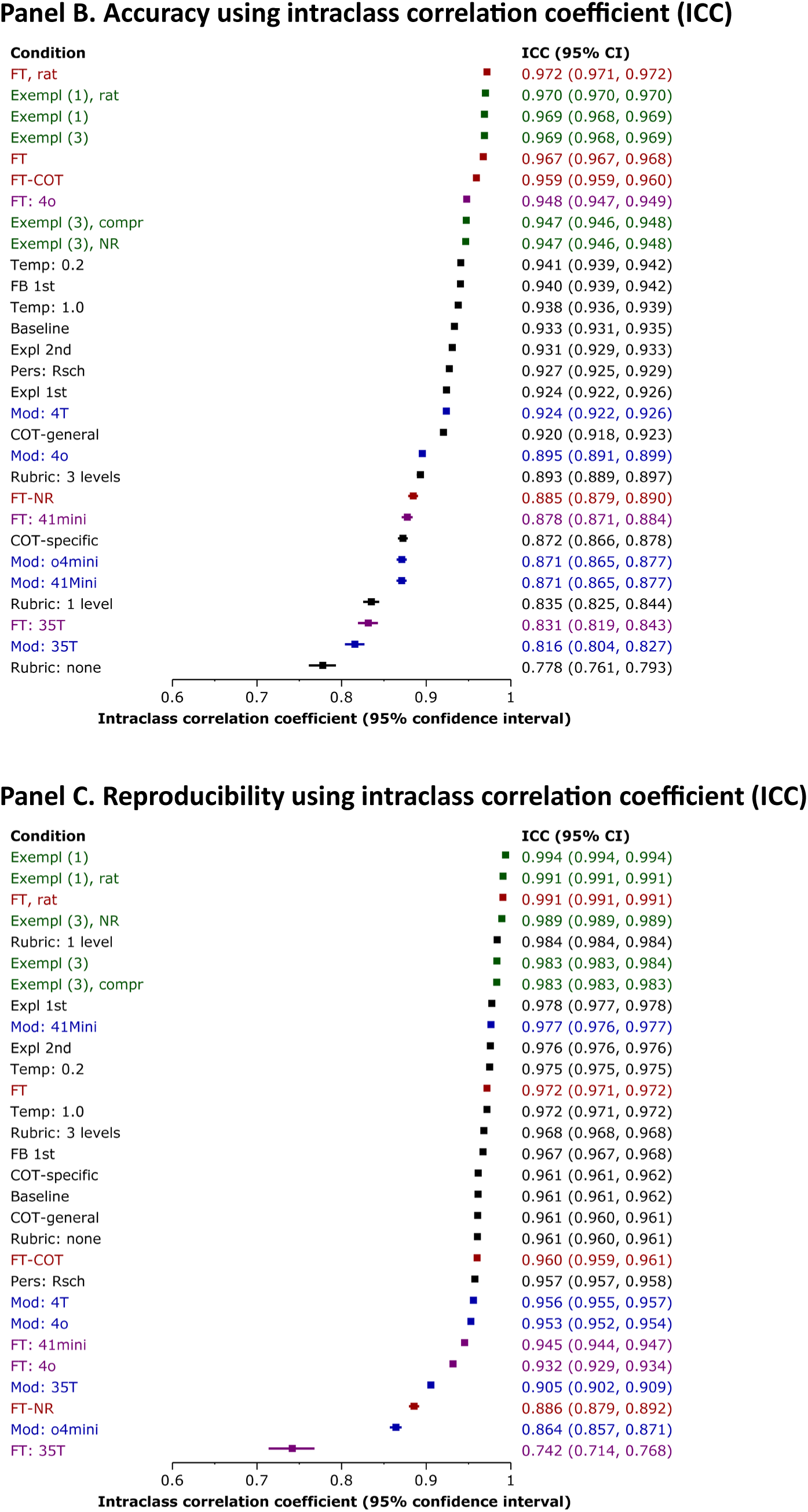
Accuracy of various prompt-model conditions used to score reflection essays using large language models. Data derive from 6 replications for each experimental condition (N=306 data points [51 essays] per condition, except for fine-tuned models N=216 data points [36 essays] per condition). See Table 1 for details on prompt-model condition variations. Colors indicate groups of related conditions that differ from baseline: green = exemplars, red = fine-tuned, blue = model other than GPT-4.1, purple = fine-tuned + other model. We calculated accuracy in comparison with human scores using mean absolute difference (MAD; Panel A) and using the single-score intraclass correlation coefficient (ICC; Panel B). We used ICC to calculate reproducibility across 6 replications using the same condition (Panel C). <u>Abbreviations</u>: FT = fine-tuned, Exempl = exemplar, rat = rationale, COT = chain-of-thought, compr = compressed essay, NR = no rubric, Temp = temperature, Expl = explanation, Mod = GPT model.

To perform formal statistical comparisons among prompt-model conditions we used the mean absolute difference (MAD) between human and GPT scores, with values of zero indicating perfect agreement. The relative accuracies (rank order) of conditions as measured using MAD and ICC were similar, although there were small differences (Figure 1, panels A and B).

The global test rejected the null hypothesis (p<.001), indicating significantly different MAD values among the prompt-model conditions. We then proceeded with pairwise comparisons of potential interest, using the Benjamini-Hochberg procedure^43^ to determine statistical significance. MAD estimates, including CIs, are detailed in Figure 1 and Table 3; we highlight selected results below.

**Table 3.** Comparisons of accuracy (mean absolute difference) among prompt-model conditions used to score reflection essays using large language models.

| Model or prompt | Sub-condition A<br>MAD<sup>a</sup> (95% CI) | Sub-condition B<br>MAD<sup>a</sup> (95% CI) | Difference<br>MAD (95% CI)<sup>b</sup> |
| --- | --- | --- | --- |
| **GPT models, not fine-tuned** | **Model A: GPT-4.1** | **Model B** |  |
| ... B = GPT-4-Turbo | 0.37 | 0.44 | -0.08 (-0.30, 0.14) |
| ... B = GPT-o4-mini | 0.37 | 0.55 | -0.19 (-0.45, 0.08) |
| ... B = GPT-4o | 0.37 | 0.63 | -0.26 (-0.50, -0.03) |
| ... B = GPT-4.1-mini | 0.37 | 0.79 | **-0.42 (-0.68, -0.17)<sup>b</sup>** |
| ... B = GPT-3.5-Turbo | 0.37 | 0.85 | **-0.48 (-0.73, -0.24)<sup>b</sup>** |
| **GPT models, fine-tuned<sup>d</sup>** | **Model A: GPT-4.1** | **Model B** |  |
| ... B = GPT-4o | 0.23 | 0.27 | -0.04 (-0.24, 0.16) |
| ... B = GPT-3.5-Turbo | 0.23 | 0.63 | **-0.40 (-0.63, -0.17)<sup>b</sup>** |
| ... B = GPT-4.1-mini | 0.23 | 0.70 | **-0.47 (-0.77, -0.17)<sup>b</sup>** |
| **Fine-tuning<sup>d</sup>** | **Fine-tuned** | **Non-fine-tuned** |  |
| All conditions | 0.45 (0.38, 0.53) | 0.70 (0.62, 0.77) | **-0.24 (-0.34, -0.14)<sup>b</sup>** |
| GPT-4.1-mini | 0.70 | 0.79 | -0.09 (-0.43, 0.24) |
| GPT-4.1 | 0.23 | 0.37 | -0.14 (-0.35, 0.07) |
| GPT-4.1 with chain-of-thought | 0.25 | 0.42 | -0.17 (-0.40, 0.06) |
| GPT-3.5-Turbo | 0.63 | 0.85 | -0.22 (-0.48, 0.04) |
| GPT-4o | 0.27 | 0.63 | **-0.36 (-0.58, -0.13)<sup>b</sup>** |
| GPT-4.1 without rubric | 0.64 | 1.12 | **-0.48 (-0.70, -0.25)<sup>b</sup>** |
| **Scoring rubric** | **More rubric** | **Less rubric** |  |
| Full (6 levels) vs<br>... Partial (3 levels) | 0.37 | 0.66 | **-0.29 (-0.53, -0.05)<sup>b</sup>** |
| ... Partial (1 level) | 0.37 | 1.01 | **-0.64 (-0.87, -0.42)<sup>b</sup>** |
| ... Partial (3 or 1 levels) | 0.37 | 0.83 (0.71, 0.95) | **-0.47 (-0.68, -0.25)<sup>b</sup>** |
| ... None | 0.37 | 1.12 | **-0.75 (-0.98, -0.52)<sup>b</sup>** |
| Any (1, 3, or 6 levels) vs None | 0.68 (0.58, 0.77) | 1.12 | **-0.44 (-0.66, -0.22)<sup>b</sup>** |
| **Few-shot learning (exemplars)** | **Exemplars** | **No exemplars** |  |
| All conditions: Exemplars = any | 0.30 (0.32, 0.37) | 0.74 (0.63, 0.86) | **-0.44 (-0.57, -0.31)<sup>b</sup>** |
| All conditions with rubric: Exemplars = any | 0.26 (0.19, 0.34) | 0.37 (0.21, 0.52) | -0.10 (-0.28, 0.07) |
| With rubric: Exemplars = 1 | 0.24 | 0.37 | -0.12 (-0.33, 0.08) |
| ... Exemplars = 3 | 0.24 | 0.37 | -0.13 (-0.33, 0.06) |
| ... Exemplars = 3 compressed | 0.31 | 0.37 | -0.05 (-0.27, 0.17) |
| Without rubric: Exemplars = 3 | 0.42 | 1.12 | **-0.70 (-0.93, -0.48)<sup>b</sup>** |
| **Justification narrative** | **Feedback** | **Rationale** |  |
| Few-shot learning: Exemplars = 1 | 0.24 | 0.24 | 0.01 (-0.17, 0.18) |
| Fine-tuned GPT-4.1<sup>d</sup> | 0.23 | 0.23 | 0.00 (-0.21, 0.21) |
| **Other hypotheses** | **Baseline<sup>c</sup>** | **Prompt B** |  |
| Persona: B = Researcher | 0.37 | 0.39 | -0.02 (-0.25, 0.21) |
| Sequence: B = Feedback first | 0.37 | 0.38 | -0.01 (-0.23, 0.21) |
| Chain-of-thought: B = General chain-of-thought | 0.37 | 0.42 | -0.06 (-0.29, 0.18) |
| Chain-of-thought: B = Specific chain-of-thought | 0.37 | 0.64 | -0.28 (-0.56, 0.00) |
| Explanation: B = Explanation first | 0.37 | 0.44 | -0.08 (-0.32, 0.17) |
| Explanation: B = Explanation second | 0.37 | 0.38 | -0.01 (-0.25, 0.23) |
| Temperature: B = Temperature low (0.2) | 0.37 | 0.34 | 0.03 (-0.20, 0.25) |
| Temperature: B = Temperature high (1.0) | 0.37 | 0.35 | 0.01 (-0.21, 0.24) |

<sup>a</sup> MAD = mean absolute difference (the difference between GPT and human scores; zero indicates perfect agreement). We report 95% confidence intervals (CIs) only for point estimates not included in Figure 1. Data derive from 6 replications for each experimental condition (N=306 data points [51 essays] per condition, except for fine-tuned models N=216 data points [36 essays] per condition).
<sup>b</sup> Bolded text indicates differences that are statistically significant using the Benjamini-Hochberg procedure.<sup>43</sup>
<sup>c</sup> Baseline prompt used GPT-4.1 with the persona of a writing evaluator, full rubric, no exemplars, numeric score before feedback, no chain-of-thought, no fine-tuning, no explanation, and temperature = 0.7 (see Table 1).

*GPT models without fine-tuning*. Accuracy varied across the models without fine-tuning, with MADs as follows: GPT-4.1, 0.37; GPT-4-Turbo, 0.44; GPT-o4-mini, 0.55; GPT-4o, 0.63; GPT-4.1-mini, 0.79; GPT-3.5-Turbo, 0.85 (see Figure 1 for 95% CIs around estimates). Pairwise comparisons were statistically significant for GPT-4.1 vs GPT-4.1-mini and GPT-3.5-Turbo (see details in Table 3).

*GPT models with fine-tuning*. Accuracy varied across the models with fine-tuning, with MADs as follows: GPT-4.1, 0.23; GPT-4o, 0.27; GPT-3.5-Turbo, 0.63; GPT-4.1-mini, 0.70. Once again, comparisons were statistically significant for GPT-4.1 vs GPT-3.5-Turbo and GPT-4.1-mini.

*Fine-tuning vs non-fine-tuning*. Differences in accuracy between fine-tuned vs non-fine-tuned models varied across conditions but always favored fine-tuning, as follows: GPT-4.1-mini, difference in MAD = – 0.09; GPT-4.1 with rubric, –0.14; GPT-3.5-Turbo, –0.22; GPT-4o, –0.36; and GPT-4.1 without rubric, –0.48. Comparisons were statistically significant for GPT-4o and GPT-4.1 without rubric. When averaging across conditions, fine-tuned models were significantly more accurate than non-fine-tuned models (mean difference –0.24 [CI, –0.34, –0.14]).

*Scoring rubric*. We found an ordinal trend of decreasing accuracy with fewer rubric levels (P<.001), with MAD 0.37, 0.66, 1.01, and 1.12 for 6 (full rubric), 3, 1, and 0 (no rubric) levels, respectively. All pairwise comparisons were statistically significant except 1 vs 0 levels (difference –0.11 [CI, –0.34, 0.13]).

*Exemplars (few-shot prompting)*. On average, conditions with exemplars (few-shot prompting) were significantly more accurate than those without (difference –0.44 [-0.57, –0.31]). This difference was largest (and statistically significant) for conditions without rubric (difference –0.70). All other pairwise comparisons favored exemplars but were not statistically significant (differences ranging –0.05 to –0.13). Accuracy for 3 exemplars (0.235) and 1 exemplar (0.242) was similar (difference –0.01 [CI, –0.18, 0.16]). Full exemplars (0.24) were more accurate than compressed exemplars (0.31), but not significantly so (difference –0.08 [CI, –0.27, 0.11]).

*Other contrasts*. None of the other hypothesized differences between prompt-model conditions reached statistical significance. Specifically, in comparison with the baseline condition (MAD = 0.37):

- Temperature made little difference, with accuracy for low and high temperatures slightly better (MAD 0.34 and 0.35, respectively) than the baseline moderate temperature.
- Asking GPT to provide feedback before the numeric score (MAD 0.38) and using a researcher persona (MAD 0.39) made little difference compared with baseline.
- In contrast with previous research using older LLMs,^18, 22, 23, 39^ explicitly directing GPT-4.1 in its “thinking” did not help. The general chain-of-thought prompt, specific chain-of-thought, and request for explanation before numeric score (MAD 0.42, 0.64, 0.44, respectively) were all worse than baseline, although not significantly so. A similar pattern emerged for the fine-tuned model (MAD for fine-tuned baseline 0.23 vs fine-tuned general chain-of-thought 0.25 [difference –0.02 [CI, –0.22, 0.18]).

Accuracies for exemplars or fine-tuning using justification narratives phrased as learner-directed feedback vs rater-directed rationales were similar (MAD 0.242 vs 0.235 for 1 exemplar, 0.230 vs 0.230 for fine-tuned).

### Within-prompt reproducibility

Within-condition reproducibility (intra-rater reliability) was “almost perfect” (ICC >0.80) for 28/29 conditions (97%), and >0.90 for 26 (90%); see Figure 1. Reproducibility ranged from 0.99 (for 1 exemplar [both feedback– and rationale-oriented justifications], fine-tuned with rationale, and 3 exemplars without rubric) to 0.74 (for fine-tuned GPT-3.5-Turbo). Distinctions among conditions were typically trivial, except for fine-tuned no rubric, GPT-o4-mini, and fine-tuned GPT-3.5-Turbo (all ICC <0.90). Notably, 5 of the top 7 most reproducible conditions involved exemplars (few-shot learning) in varying forms.

### Balancing accuracy, reproducibility, and cost

Figure 2 superimposes the outcomes of accuracy, reproducibility, and cost. Although fine-tuned models were among the most accurate and reproducible, they were also the most expensive when rating few essays. Most prompts with non-fine-tuned GPT-4.1 provided performance only slightly inferior, at much lower cost. By contrast, when rating many essays, the fine-tuned model with baseline prompt was slightly less expensive than the non-fine-tuned model.

**Figure 2.**
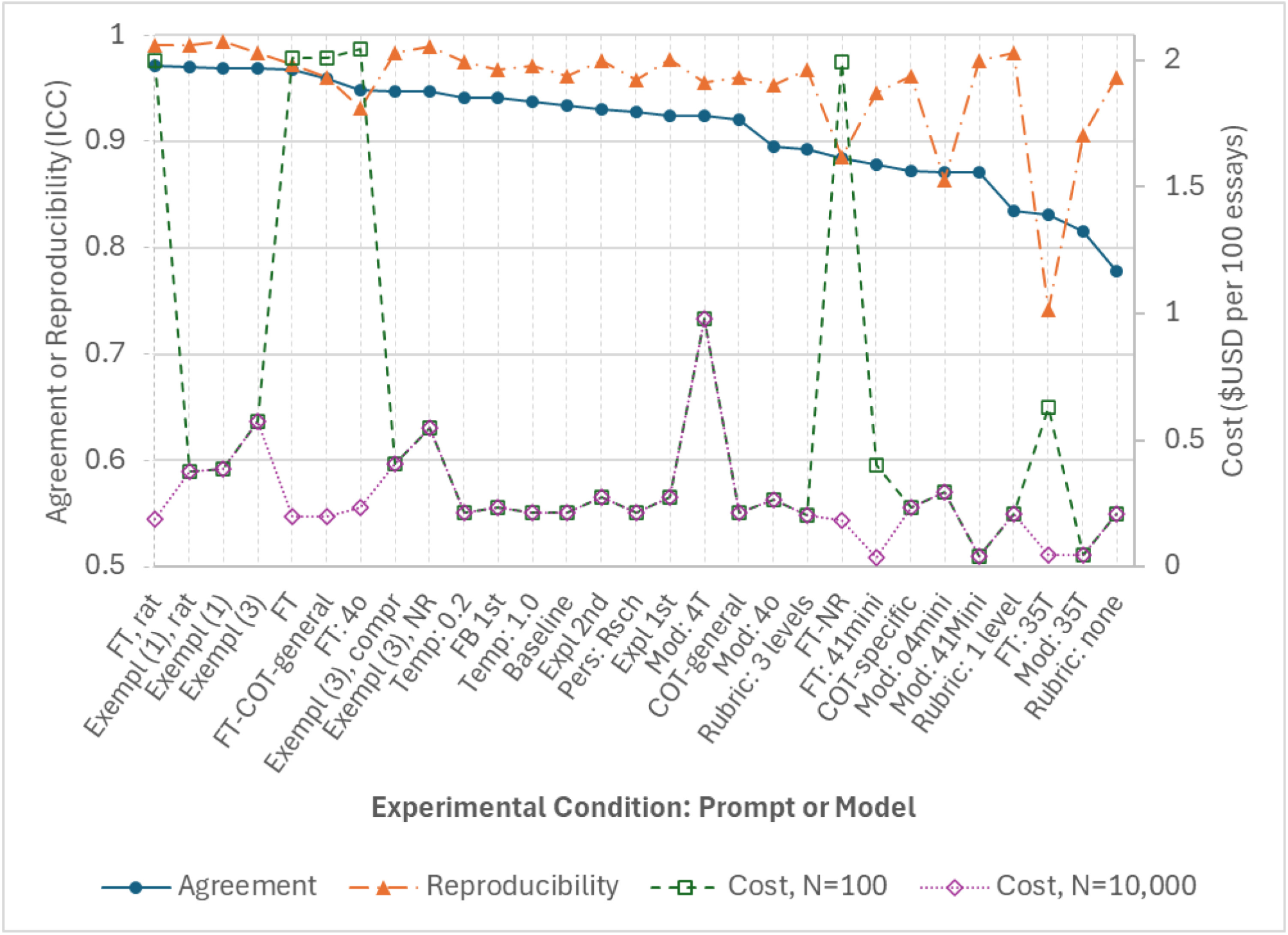
Accuracy, within-prompt reproducibility, and cost of various prompt-model conditions used to score reflection essays using large language models. Data derive from 6 replications for each experimental condition (N=306 data points [51 essays] per condition, except for fine-tuned models N=216 data points [36 essays] per condition). See Table 1 for details on prompt-model condition variations. We calculated accuracy in comparison with human scores using the single-score intraclass correlation coefficient (ICC). We calculated reproducibility as consistency across 6 replications using the same condition. We report cost for 2 situations: scoring N=100 essays and scoring N=10,000 essays. To facilitate comparison, we report the cost for both situations as standardized to 100 essays. Fine-tuned models include the one-time cost of fine-tuning, which when amortized across 10,000 essays results in a lower per-essay cost. <u>Abbreviations</u>: FT = fine-tuned, Exempl = exemplar, rat = rationale, COT = chain-of-thought, compr = compressed essay, NR = no rubric, Temp = temperature, Expl = explanation, Mod = GPT model.

## Discussion

This study investigated the accuracy, reproducibility, and cost of various LLM prompts and models for scoring medical student essays. We found overall excellent accuracy and reproducibility (ICC “almost perfect” [>0.80] for 28 of 29 conditions, for both outcomes), with costs as low as USD $0.04 per 100 essays. Fine-tuned models were more accurate than non-fine-tuned models, and prompts with exemplars (few-shot learning) and more anchors in the scoring rubric were more accurate than those without exemplars and fewer anchors. However, fine-tuned models and prompts with exemplars were also the most costly, although the per-essay cost of fine-tuned models decreased at higher essay volumes. We also used LLMs to generate 36 essays new for analysis.

### Limitations

Perhaps the most salient limitation is the use of fabricated essays, which may have better grammar and coherence than real essays and thus could inflate observed accuracy. Replication using only authentic student essays would enhance the generalizability of results. Additionally, we used only 1 LLM vendor (OpenAI); prompts may perform differently for other LLMs (e.g., Gemini, Anthropic). This study reports only monetary costs for LLM use; we do not know the costs (time or money) required for human scoring. A more detailed cost analysis would enable insightful comparisons in future research. Our findings may become dated as this field rapidly advances, yet they highlight several important implications that advance the field *now*. Although human ratings of narrative assessments are fallible, the blinded human raters had high agreement in establishing reference-standard scores. Strengths include the use of a separate LLM (Gemini) to generate new essays; hypothesis-driven comparisons of numerous state-of-the-art prompt-model conditions; and robust statistical analyses with transparent reporting.

### Integration with prior work

This study extends prior research on LLM prompt engineering for scoring narrative assessments,^18, 22, 23, 36–41, 45, 46^ which in most instances used now-outdated LLMs. These studies also often used rudimentary or flawed research designs and statistical analyses. We used that prior work to identify salient prompt variations and studied these using newer, more powerful LLMs, rigorous methods, and robust statistical analyses. Our findings also parallel research in questionnaire design that shows improved performance when more rubric anchors are provided.^47–49^

### Implications: Balancing cost and performance

The conditions yielding highest accuracy (few-shot learning and fine-tuning) were also the most expensive. The optimal conditions also depend on the number of essays to be scored. For 100 essays, if we assume that all conditions with both accuracy ICC ≥0.90 and reproducibility ICC ≥0.90 are equally acceptable, the low-temperature prompt is the best option ($0.211 per 100) and baseline is close behind ($0.212 per 100). However, for 10,000 essays, the fine-tuned rationale condition is cheaper ($0.189 per 100, saving $2.27 across all 10,000 essays), and fine-tuned baseline is close behind ($0.200 per 100). If educators are willing to accept slightly lower performance, GPT-4.1-mini had much lower cost ($0.040 per 100, saving $17.13 across 10,000 essays compared with non-fine-tuned baseline), and accuracy and reproducibility were still almost perfect (ICC ≥0.87). By contrast, few-shot prompts were more accurate but more expensive (starting at $0.376 per 100, which is $16.43 more than non-fine-tuned baseline across 10,000 essays). This finding supports assertions from LLM vendors that smaller contemporary models are “an attractive model for many use cases.”^50^

### Additional implications

Most importantly, we found very high accuracy and reproducibility when using LLMs to score medical student reflection essays using a previously-defined rubric.^42^ Our findings offer strong support for using LLMs for this task, with appropriate human supervision. LLM-supported scoring could substantially reduce educator burden and comes at reasonable cost (as little as USD $0.04 per 100 essays). With suitable rubrics, reference-standard exemplars, scoring justifications, and model training, this approach could likely be extended to other narrative assessments. Moreover, the mistakes and biases that LLMs introduce when rating may be different than those introduced by humans,^21^ and thus could complement one another.

Current LLMs were clearly superior to older models (GPT-4-Turbo, GPT-3.5-Turbo) in accuracy, cost, or both. This calls into question previous research using older LLMs. Moreover, some of our findings unexpectedly contradict previous research on prompt engineering (e.g., the lack of improvement using chain-of-thought and explanation-first prompts^18, 22, 23, 39^), and align with a recent study showing similar lack of effect.^51^ Further investigation would be needed to determine whether our findings reflect advancements in LLM technology or specifics of the rating task or prompt wording. Our findings indicate that updated research using the latest LLMs is essential, and likely a continuously-moving target.

We note that o4-mini (a deep-reasoning LLM) and prompts requesting chain-of-thought or explanation (i.e., additional LLM reasoning) were the slowest conditions (>4.5 sec per essay). Surprisingly, these high-reasoning conditions demonstrated slightly *worse* accuracy than the baseline prompt.

Beyond their use in scoring, this study illustrates the use of LLMs to generate new data (36 new essays [Appendix Table A1] and 3 compressed exemplars [Appendix Box 2]) to support research. AI-created data have been previously used in research.^22, 40, 41, 45^ We also used LLMs for support in developing prompts (see Appendix Box 2) and in writing the Python code. In all cases, we remained intimately involved, revising and approving the final product; but AI support substantially alleviated the human workload.

Finally, while we did find differences among prompts, in most cases the differences were small and likely of trivial consequence. Moreover, newer GPT models were all reasonably accurate and reproducible for all tested prompt conditions, and the near-perfect ICCs leave little room for improvement. This suggests that current LLMs may not require the same sophistication and nuance in prompt engineering as earlier models. The generalizability of these findings to other narrative assessment tasks (other essay topics, post-encounter clinical notes, clinician-patient conversations, and ratings of feedback quality) and to LLMs other than OpenAI’s GPTs warrants further investigation. For now, our findings suggest that educators can adapt our baseline prompt to their to specific purpose, and use this with GPT-4.1 or 4.1-mini (or, we surmise, another state-of-the-art LLM) to score student reflection essays with high accuracy and reproducibility. For large-volume tasks, fine-tuned models offer best performance and lowest cost. When rating few essays, a single exemplar offers best performance, but the baseline prompt without exemplars was nearly as good and much less expensive.

## Acknowledgements

The authors thank Patricia O’Sullivan for permission to use the scoring rubric and associated essays. Computer tools (including OpenAI GPT and Gemini large language models) were used to analyze essays, generate new essays and compressed exemplars, draft prompts, and help write Python code. They played no role in the writing of the manuscript itself.

## Ethical approval

This study did not involve human subjects.

## Disclaimers and authorship

Author DAC affirms that the authors had access to all the study data, take responsibility for the accuracy of the analysis, and had authority over manuscript preparation and the decision to submit the manuscript for publication. All authors approved the final manuscript and agree to adhere to all terms of authorship and copyright.

## Data availability

The verbatim prompt for each prompt-model condition is specified in Table 1. The custom Python script used to access the OpenAI API was designed specifically for internal data processing and research rigor. This script was developed as a task-specific research tool and is not currently packaged for public distribution.

## Appendices

**Appendix Box 1.**
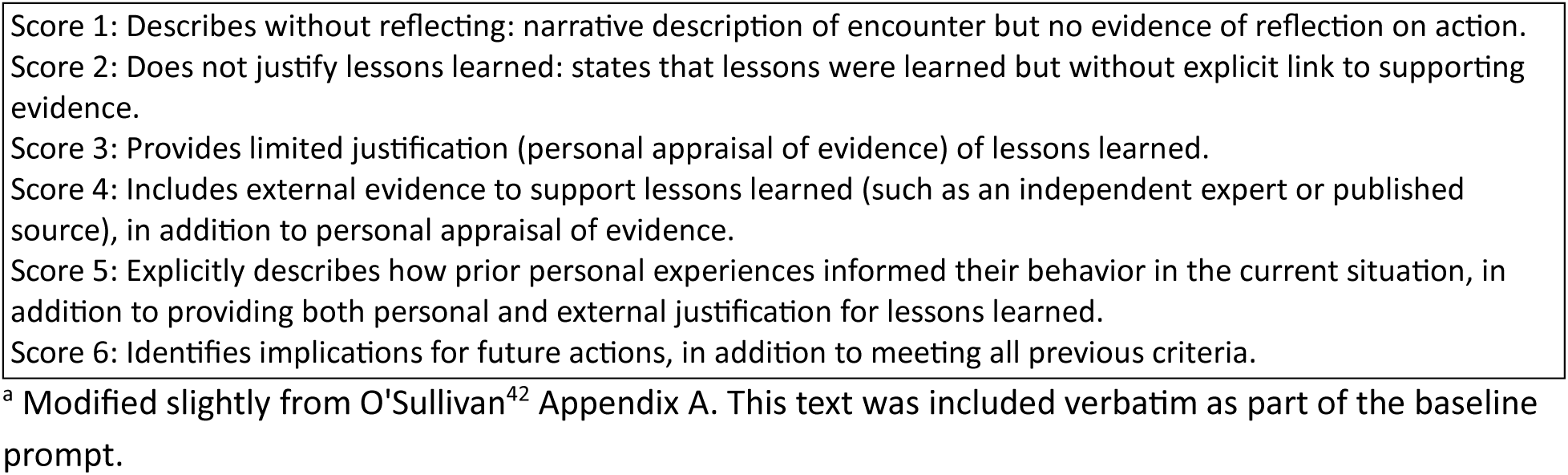
Reflection scoring rubric^a^.

|  |
| --- |
| Score 1: Describes without reflecting: narrative description of encounter but no evidence of reflection on action.<br>Score 2: Does not justify lessons learned: states that lessons were learned but without explicit link to supporting evidence.<br>Score 3: Provides limited justification (personal appraisal of evidence) of lessons learned.<br>Score 4: Includes external evidence to support lessons learned (such as an independent expert or published source), in addition to personal appraisal of evidence.<br>Score 5: Explicitly describes how prior personal experiences informed their behavior in the current situation, in addition to providing both personal and external justification for lessons learned.<br>Score 6: Identifies implications for future actions, in addition to meeting all previous criteria. |

<sup>a</sup> Modified slightly from O'Sullivan<sup>42</sup> Appendix A. This text was included verbatim as part of the baseline prompt.

**Appendix Box 2.**
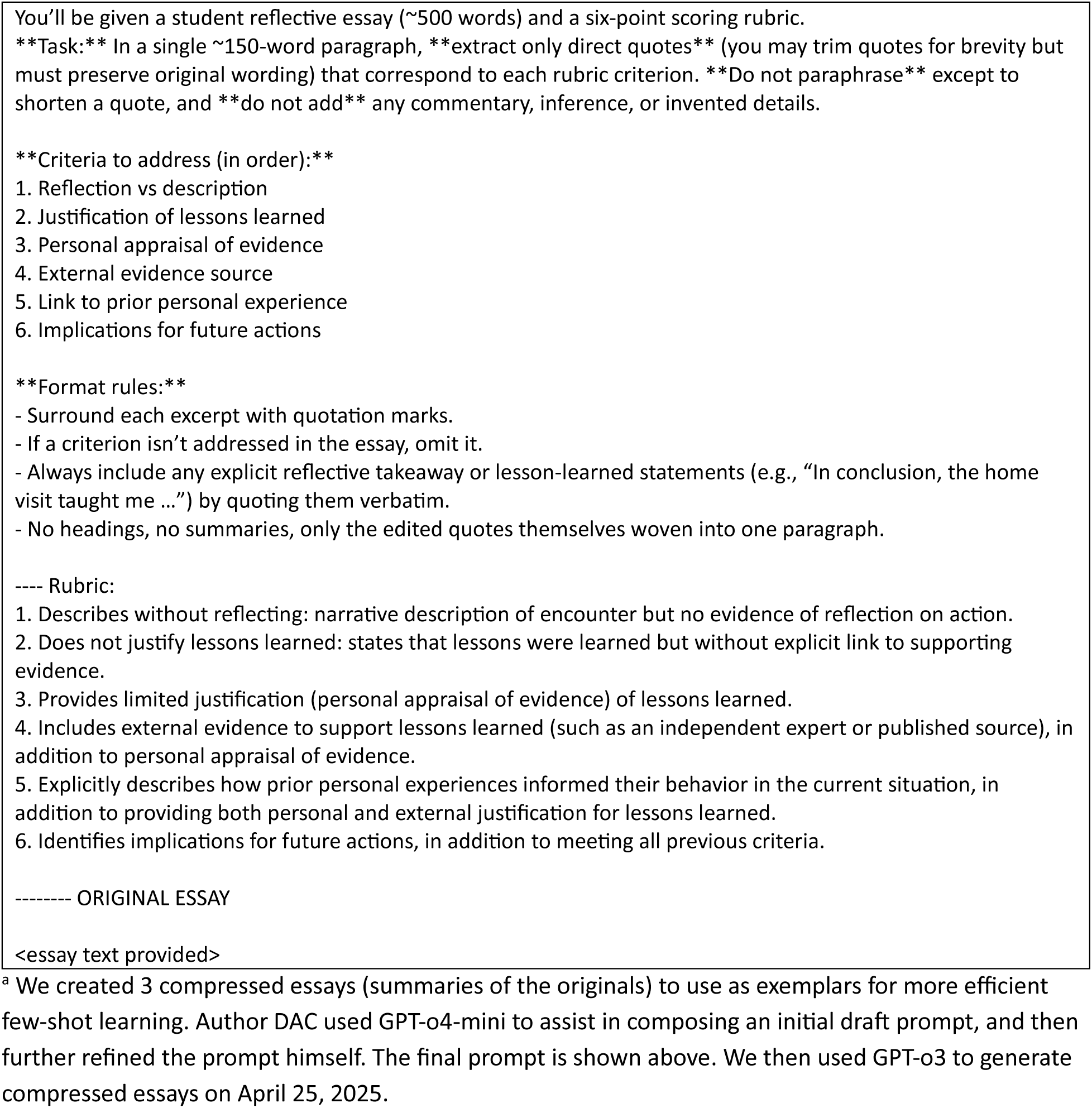
Prompt used with GPT-o3 to generate 3 compressed exemplar reflection essays^a^.

**Appendix Table A1.** Prompting dialog used with Gemini to generate 36 new reflection essays^a^.

| Prompt | Gemini response |
| --- | --- |
| **Initial Generation** |  |
| I want you to help me generate new data for a research project. I have 18 reflective essays written by medical students. I want you to generate a new set of reflective essays that are similar in their writing quality to the original 18. The idea is that the essays will range in quality from no reflection to extremely high reflection. I will upload the scoring rubric to help you norm your essay writing. Please be creative in the essay content - I want you to vary the medical specialty (medicine, surgery, psychiatry, pediatrics, etc) and the patient (man, woman, age) and clinical condition (anything you want). However, I want the depth of reflection to match each example essay. You will generate 18 new essays with a 1-to-1 match for style, quality, and numeric score.<br>Here is the prompt that students responded to: Reflect on a recent clinical or other professional situation where you made a mistake or didn't have the necessary knowledge or skills. What did you learn from this experience? What information supports your conclusions about lessons learned? How will this affect your future actions?<br>Let's think thru this before we start. Please explain the task and how you will approach it. | Okay, I understand the task. You want me to generate 18 new reflective essays written from the perspective of medical students, mirroring the style, quality, and depth of reflection present in your original 18 essays. These new essays should also address the same prompt you provided to your students.<br>Here's a breakdown of the task and my approach:<br>Task:<br>Generate 18 new reflective essays.<br>Match each new essay to one of your original 18 essays in terms of:<br>Writing Quality: Sentence structure, vocabulary, organization, and overall clarity.<br>Depth of Reflection: This is the most crucial aspect. The new essay should exhibit a similar level of self-awareness, critical analysis of the experience, identification of learning points, and articulation of how future actions will be influenced. This will be guided by the scoring rubric you will provide.<br>Numeric Score: Although I won't be able to assign a score without the rubric, my goal is to create essays that would likely receive a similar score to their corresponding original essay based on the depth of reflection and writing quality.<br>Style: The overall tone and approach of the writing.<br>Vary the content of the new essays by:<br>Medical Specialty: Each essay will focus on a different area of medicine (e.g., internal medicine, general surgery, psychiatry, pediatrics, obstetrics and gynecology, emergency medicine, etc.).<br>Patient Demographics: The essays will involve diverse patients (man, woman, child, different ages).<br>Clinical Condition: The scenarios will involve a wide range of medical issues and situations.<br>Address the original prompt: Each new essay will reflect on a situation where the student made a mistake or lacked knowledge/skills, the lessons learned, supporting information, and the impact on future actions.<br>My Approach: |
|  | Awaiting the Rubric: The first and most critical step is for you to provide the scoring rubric. This rubric will be my guide for understanding what constitutes different levels of reflection and writing quality. I will carefully analyze the rubric to identify the key criteria for each score level.<br>Norming with Original Essays (Implicit): Once you provide the rubric, I will implicitly "norm" my understanding of the scoring levels by comparing them to the characteristics I can infer from your description of the original 18 essays (ranging from no reflection to extremely high reflection). While I won't see the original essays directly, your description helps me understand the spectrum I need to create.<br>Generating New Scenarios: For each of the 18 new essays, I will create a unique scenario involving a different medical specialty, patient, and clinical condition. These scenarios will be designed to naturally lend themselves to different levels of reflection, mirroring the expected depth in the corresponding original essay.<br>Matching Depth of Reflection: This will be the core of the generation process. For each new essay, I will aim for a specific level of reflection based on my understanding of the rubric and the implied level of reflection in its counterpart. This will involve:<br>No/Low Reflection: Focusing on a superficial description of the event without significant analysis of personal contributions, underlying factors, or concrete learning. The "lessons learned" might be vague or generic. The impact on future actions might be absent or poorly articulated.<br>Moderate Reflection: Demonstrating some awareness of personal involvement, identifying specific knowledge or skill gaps, and articulating some learning points. The connection between the experience and future actions will be present but might lack detailed explanation or supporting reasoning.<br>High/Extremely High Reflection: Showing deep self-awareness, critically analyzing the situation, identifying root causes and contributing factors, articulating specific and insightful learning points supported by reasoning, and providing a clear and detailed plan for how the experience will shape future practice.<br>Matching Writing Quality and Style: While focusing on the depth of reflection, I will also pay attention to the overall writing quality. I will aim for clear and coherent prose, appropriate vocabulary for a medical student, and a style that is consistent within each essay and broadly similar to what one might expect from medical student writing. |
|  | Addressing All Parts of the Prompt: Each essay will explicitly address all aspects of the prompt: the mistake or lack of knowledge/skills, the lessons learned, supporting information for those conclusions, and the impact on future actions.<br>Iterative Refinement (Internal): Although you won't see this, my internal process will involve some level of self-correction and refinement as I generate each essay to ensure it meets the desired criteria for reflection, writing quality, and content variation.<br>In summary, my process will be to:<br>Receive and thoroughly understand the scoring rubric.<br>Create 18 distinct clinical scenarios with varied specialties, patients, and conditions.<br>For each scenario, generate a reflective essay that mirrors the implied depth of reflection and writing quality of one of your original essays, as guided by the rubric.<br>Ensure each essay fully addresses the prompt and demonstrates a clear connection between the experience, learning, and future actions.<br>Maintain appropriate writing quality and style for a medical student.<br>I am ready to proceed once you provide the scoring rubric. I look forward to helping you with your research! |
| Not quite correct - your step 2 is wrong. I will supply 18 actual essays, and you will be able to see these. I actually want you to look at each essay, 1 by 1, and create a matching essay that is similar in quality but different in content. ....<br>Please describe again the approach you will take, so I can approve all steps. | You are absolutely correct! My apologies for the misunderstanding in step 2. Thank you for the clarification.<br>Here is the revised approach I will take, incorporating the fact that you will provide the 18 original essays for me to analyze directly:<br>Revised Approach:<br>Receive and Analyze the Original Essays: You will provide me with the 18 original reflective essays. My first step will be to carefully read and analyze each essay individually. For each original essay, I will focus on identifying:<br>Depth of Reflection: I will use the scoring rubric you provide to assess the level of reflection demonstrated in the essay. This will involve identifying the extent of self-awareness, critical analysis, articulation of learning, and future application.<br>Writing Quality: I will evaluate the essay's clarity, organization, sentence structure, vocabulary, and overall coherence. |
|  | Style and Tone: I will note the writing style, including the formality, personal voice, and any specific stylistic elements.<br>Content Characteristics (as a reference point, but the new content will be different): I will briefly note the medical specialty, patient type, and clinical situation described to ensure the new essay has entirely different content.<br>Overall Impression: I will form a holistic understanding of the essay's strengths and weaknesses in relation to the prompt and the expected qualities of reflective writing.<br>Generate a New, Matching Essay (One-to-One): For each of the 18 original essays, I will generate a new reflective essay that aims to match it in the following aspects:<br>Depth of Reflection: The new essay will demonstrate a level of reflection that closely aligns with the level observed in its corresponding original essay, as determined by the scoring rubric. If the original essay shows minimal reflection, the new one will also reflect at a similar low level. Conversely, a highly reflective original essay will be matched with a new essay demonstrating a similarly high level of insight and analysis.<br>Writing Quality: The new essay will be written with a similar level of clarity, organization, and grammatical correctness as its corresponding original essay. I will aim for comparable sentence complexity and vocabulary.<br>Style and Tone: I will attempt to capture a similar tone and writing style in the new essay.<br>Different Content: Crucially, the new essay will feature entirely different content. This will involve:<br>A different medical specialty (e.g., if the original was about surgery, the new one might be about psychiatry).<br>A different patient (e.g., different age, gender, background).<br>A different clinical condition or professional situation.<br>A new and distinct mistake or lack of knowledge/skills as the focus of the reflection.<br>New and distinct lessons learned, supporting information, and future actions.<br>Review and Refine: After generating each new essay, I will briefly review it to ensure that it adequately matches the corresponding original essay in terms of the identified characteristics (depth of reflection, writing quality, and style) while presenting entirely new and varied content that addresses the original prompt.<br>In summary, my revised approach is:<br>Receive and thoroughly analyze each of the 18 original essays individually, using the provided scoring rubric to assess the depth of reflection, and also noting writing quality and style. |
|  | For each original essay, generate a new reflective essay that mirrors its corresponding original in depth of reflection, writing quality, and style, but features entirely different content (medical specialty, patient, condition, mistake, learning, and future actions) while still addressing the original prompt.<br>Briefly review each new essay to ensure it meets the criteria.<br>Please let me know if this revised approach is accurate and meets your requirements. I am ready to proceed once you provide the original essays and the scoring rubric. |
| Sounds good. Here is the scoring rubric. Here is a file with all essays. | Before I begin generating the new essays, I just want to confirm a few things to ensure I'm on the right track:<br>Rubric Understanding: I have carefully reviewed the rubric. I understand the criteria for each score level (1-6) and the key differentiators between them, particularly the emphasis on the use of external evidence for higher scores and the progression from description to analysis and integration for the highest scores.<br>Essay Analysis: I have read each of the 18 essays and noted the assigned numeric score for each. |
| [After generating 36 new essays, we noted that those at level 1 and 2 were rather good, and those at level 5 were not very different from those at level 4. The level 1 essays were also abnormally brief. We thus proceeded with manual correction for essays at these levels, using prompts below.] |  |
| **Manual correction: Level 1** |  |
| Thanks. As I review these, there are not very many that are poor quality. The essays you've written are too good! ... I am going to give you the text of an essay, and I want you to rewrite it to make it LOWER quality. The grammar and style can remain the same, but I want you to make it NOT reflective. I want it to meet the following criteria:<br>Describes without reflecting. Narrative description of encounter but no evidence of reflection on action. | <Revised essay text returned.> |
| I also want you to add a few additional details to the story (without reflection) so that the length is approximately the same as the original (within 20% word count).<br>As you do the rewrite, make sure to make this an INTERESTING experience. It should be an experience that would be potentially important or transformative; but the essay just doesn't show any reflection on that transformative potential.<br><Essay text provided> |  |
| **Manual correction: Level 2** |  |
| Now let's shift gears. We are going to do the same thing, but this time we will retain a LITTLE BIT of reflection. The essay will identify 1 or 2 vague lessons learned, but will not clearly link these lessons to the story. It will be very poor reflection, very vague. Here are the specific instructions for this level of reflection: "Does not justify lessons learned. States that lessons were learned but without explicit linkage to supporting evidence. Vague reference to lessons learned without elaboration. List of lessons learned without linkage to evidence. General platitudes about optimal care without specific linkage to scenario."<br><Essay text provided> | <Revised essay text returned.> |
| **Manual correction: Level 5** |  |
| Now we will aim for level 5. ... The original essays I will give you are not good enough - they need to be improved. In particular, they lack external evidence; this could be "detailed feedback from patients or professional associates, objective | Okay, I understand the task. I will take the original essay you provide and rewrite it to meet the criteria for a level 5 reflection. This means I will:<br>Retain the elements of a level 4 reflection (including external evidence). |
| data on outcomes, and/or use of the literature." However, we need to REMOVE any reference to future plans or implications. It would also be good if they were a little longer. here you go. ... Tell me what you are going to do, and then I'll give you the first essay. | Add analysis of factors from the experience, explicitly referring to prior experiences and describing how they inform the narrator's behavior in the current situation.<br>Ensure that all references to future plans or implications are removed.<br>Essentially, I will deepen the reflection by connecting it to the narrator's past experiences and showing how those experiences shaped their actions and understanding of the situation, all while backing up the reflection with external evidence. |
| Very good, but can you please add a lesson learned (without specifically mentioning the future).<br><Essay text provided> | <Revised essay text returned.> |

<sup>a</sup> We used the large language model, Gemini-2.0-flash, to create 36 new essays modeled after the original 18 published essays, on April 18-19, 2025. This table summarizes the interaction between author DAC and Gemini.

**Appendix Table A2.**
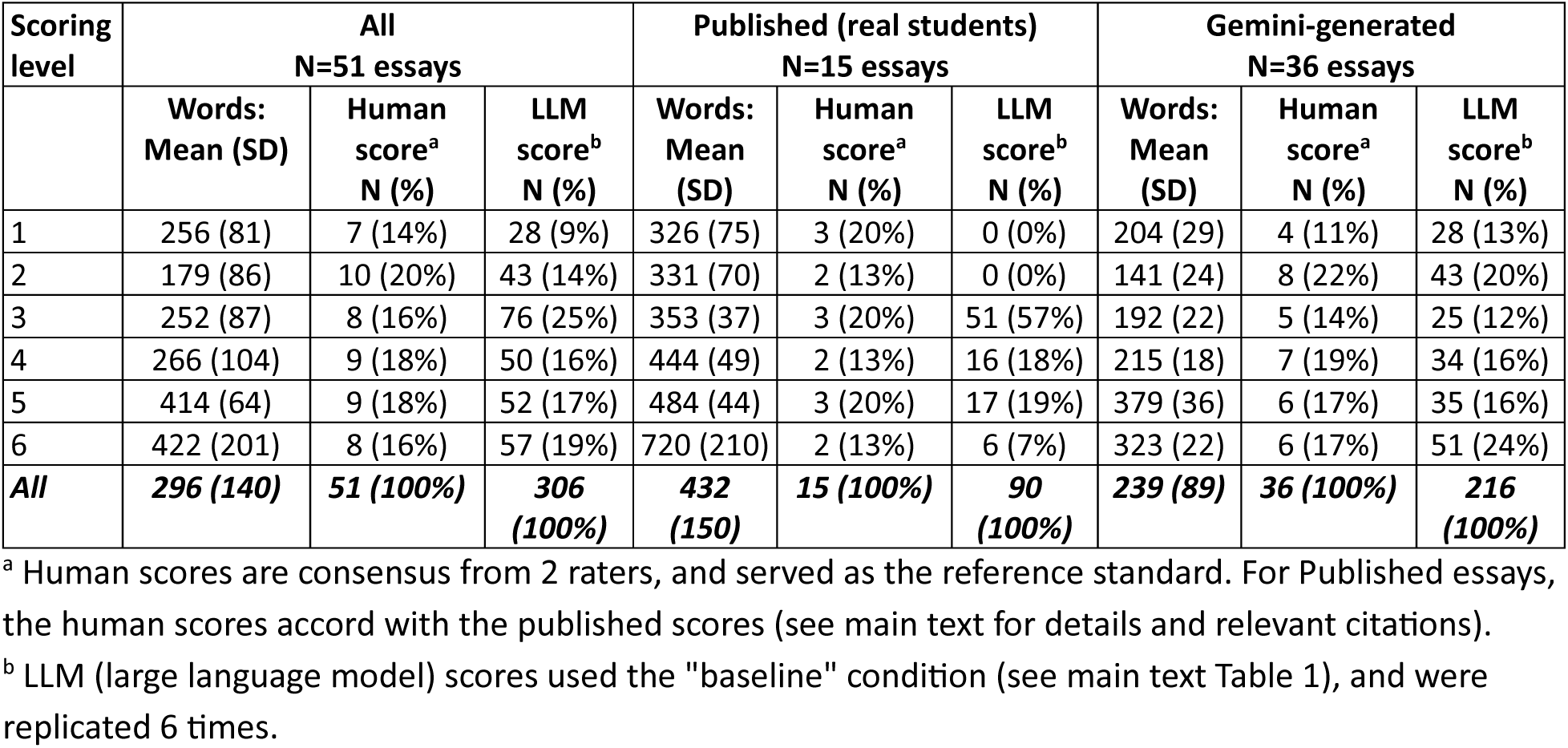
Length and score distribution of reflection essays scored by large language models.

| Scoring level | All<br>N=51 essays |  |  | Published (real students)<br>N=15 essays |  |  | Gemini-generated<br>N=36 essays |  |  |
| --- | --- | --- | --- | --- | --- | --- | --- | --- | --- |
|  | Words:<br>Mean (SD) | Human<br>score<sup>a</sup><br>N (%) | LLM<br>score<sup>b</sup><br>N (%) | Words:<br>Mean<br>(SD) | Human<br>score<sup>a</sup><br>N (%) | LLM<br>score<sup>b</sup><br>N (%) | Words:<br>Mean<br>(SD) | Human<br>score<sup>a</sup><br>N (%) | LLM<br>score<sup>b</sup><br>N (%) |
| 1 | 256 (81) | 7 (14%) | 28 (9%) | 326 (75) | 3 (20%) | 0 (0%) | 204 (29) | 4 (11%) | 28 (13%) |
| 2 | 179 (86) | 10 (20%) | 43 (14%) | 331 (70) | 2 (13%) | 0 (0%) | 141 (24) | 8 (22%) | 43 (20%) |
| 3 | 252 (87) | 8 (16%) | 76 (25%) | 353 (37) | 3 (20%) | 51 (57%) | 192 (22) | 5 (14%) | 25 (12%) |
| 4 | 266 (104) | 9 (18%) | 50 (16%) | 444 (49) | 2 (13%) | 16 (18%) | 215 (18) | 7 (19%) | 34 (16%) |
| 5 | 414 (64) | 9 (18%) | 52 (17%) | 484 (44) | 3 (20%) | 17 (19%) | 379 (36) | 6 (17%) | 35 (16%) |
| 6 | 422 (201) | 8 (16%) | 57 (19%) | 720 (210) | 2 (13%) | 6 (7%) | 323 (22) | 6 (17%) | 51 (24%) |
| **All** | **296 (140)** | **51 (100%)** | **306 (100%)** | **432 (150)** | **15 (100%)** | **90 (100%)** | **239 (89)** | **36 (100%)** | **216 (100%)** |

<sup>a</sup> Human scores are consensus from 2 raters, and served as the reference standard. For Published essays, the human scores accord with the published scores (see main text for details and relevant citations).
<sup>b</sup> LLM (large language model) scores used the "baseline" condition (see main text Table 1), and were replicated 6 times.

**Appendix Table A3.**
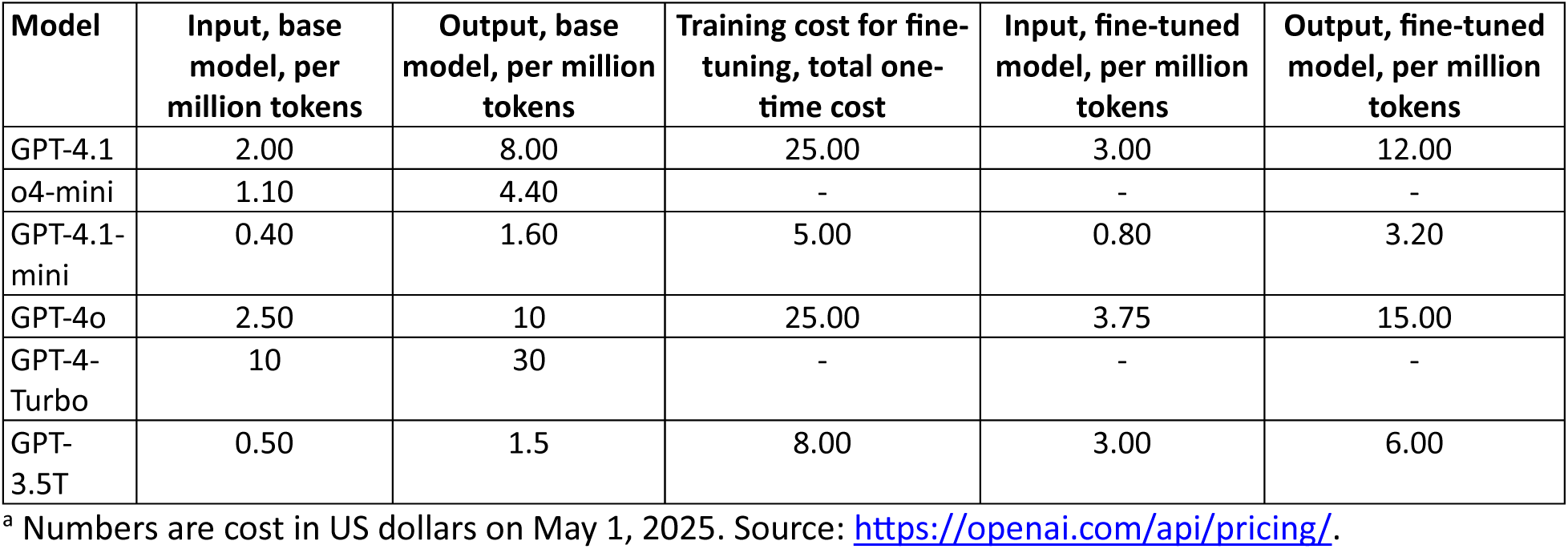
Per-token pricing used to calculate the cost of scoring reflection essays with large language models ^a^.

